# Spatial Analysis Uncovers Immune Resistance Mechanisms in Non-Beneficial Hepatocellular Carcinoma Treated with Y90 Radioembolization-Nivolumab

**DOI:** 10.64898/2026.07.10.26357712

**Authors:** Mai Chan Lau, Denise Goh, Marcia Zhang, Menaka P. Rajapakse, Wei Kit Tan, Zhen Yuan Chew, Xing Yi Woo, Zhen Wei Neo, Xinru Lim, Ye Jiangfeng, Zhu Zhu, Zhengyang Wang, Juha P. Väyrynen, David Tai, Joe Yeong

## Abstract

**Background & Aims:** Hepatocellular carcinoma (HCC) remains a leading cause of cancer mortality, with most patients presenting at advanced stages requiring systemic therapy. Despite promising outcomes with immune checkpoint inhibitors (ICI), responses remain variable due to an immunosuppressive tumor microenvironment. Y90 radioembolization offers potential immune priming, but only a subset of patients benefit. Here, we apply spatial multi-omics to delineate baseline and treatment-induced immune features and identify predictive signatures of progressive disease (PD) for early detection of patients unlikely to benefit from therapy.

**Approach & Results:** Paired baseline (Day 0) and on-treatment (Day 35) biopsies were obtained from 33 patients, following Y90 radioembolization (Day 14) and nivolumab. Multiplex immunohistochemistry (mIHC) was used for cell–cell interaction analysis. A subset was further profiled using Visium (n=13) for tissue category–specific analysis and NanoString GeoMx DSP (n=12) for cell type–resolved transcriptomic and pathway analyses. Global spatial transcriptomics analysis revealed minimal baseline immune activity in PD, indicating an intrinsically immune-deficient TME. Despite treatment-induced activation, PD exhibited reduced CD8⁺ T cell abundance and limited reinvigoration of exhausted subsets, and persistent LAG-3–associated exhaustion. DSP showed downregulation of antigen presentation and T cell activation pathways. Macrophage profiling revealed enrichment of CD38⁺ phenotypes, contrasting CXCL9–CXCR3–associated responses in responders. Furthermore, a 72-gene PD signature was identified and validated in TCGA, associating with poorer survival.

**Conclusions:** Integrated spatial multi-omics reveals that PD in HCC is associated with an immune-deficient TME, characterized by LAG-3–associated CD8⁺ exhaustion and immunosuppressive macrophages. A 72-gene signature enables early identification and supports alternative therapeutic strategies.

## Introduction

Hepatocellular carcinoma (HCC) is often diagnosed at an advanced stage, where curative interventions including surgical resection and liver transplantation are often restricted by underlying liver dysfunction or the extent of the tumor(1). Systemic therapies have become the mainstay and immunotherapy-based combination strategies have emerged as alternative therapeutic options. However, majority of HCC are immunologically ‘cold’ with an immunosuppressive tumor microenvironment (TME)—characterised by low immune infiltration—that do not respond to immune checkpoint inhibition (ICI) alone(2). As such, radioembolization with yttrium-90 (Y90RE) has emerged as a promising approach that not only delivers locoregional control but also re-activate the immune system and overcome resistance(3, 4). The combination of Y90RE with Nivolumab (Y90RE-Nivolumab) yielded promising results in therapeutic synergism and safety profile, where the objective response rates ranged from 30.6 to 41.5%(5, 6). It underscores the need for in-depth TME profiling to better understand clinical outcomes and to delineate distinct immunological contributions that underlies treatment responsiveness.

Emerging evidence suggests that both inherent features of the TME and treatment-induced immune modulation play a role in shaping clinical outcomes(7, 8). Inherent features may include tumor immunogenicity, baseline levels of immune cell infiltration, and expression of immune checkpoints molecules. Hence in this study, we applied spatial multi-omics techniques to comprehensively profile the TME of advanced HCC. Specifically, we aimed to identify inherent (baseline) and treatment-induced (on-treatment) features that may contribute to treatment non-responsiveness. We also sought to identify a predictive signature associated with progressive disease to enable early identification of patients unlikely to benefit from therapy and thereby avoid unnecessary ineffective treatment.

## Methods

### Sample Collection

For each of the 33 patients with documented treatment response (see Supplementary Information), paired baseline (Day 0) and on-treatment (Day 35) biopsies were obtained following Y90RE (Day 14) and one dose of intravenous nivolumab(9). All 33 paired tissue samples underwent multiplexed immunohistochemistry (mIHC), while subsets were profiled using 10x Visium (n=13) and NanoString GeoMx Digital Spatial Profiler (DSP, n=12) (**Fig. 1A**). The ethical approval for this study was provided by the Agency for Science, Technology and Research (A*STAR), IRB: 2021-112 and by SingHealth Centralised Institutional Review Board IRB Ref. No. 2016/2613. The study was performed in accordance with the Declaration of Helsinki and all patients provided written consent.

**Fig. 1.**
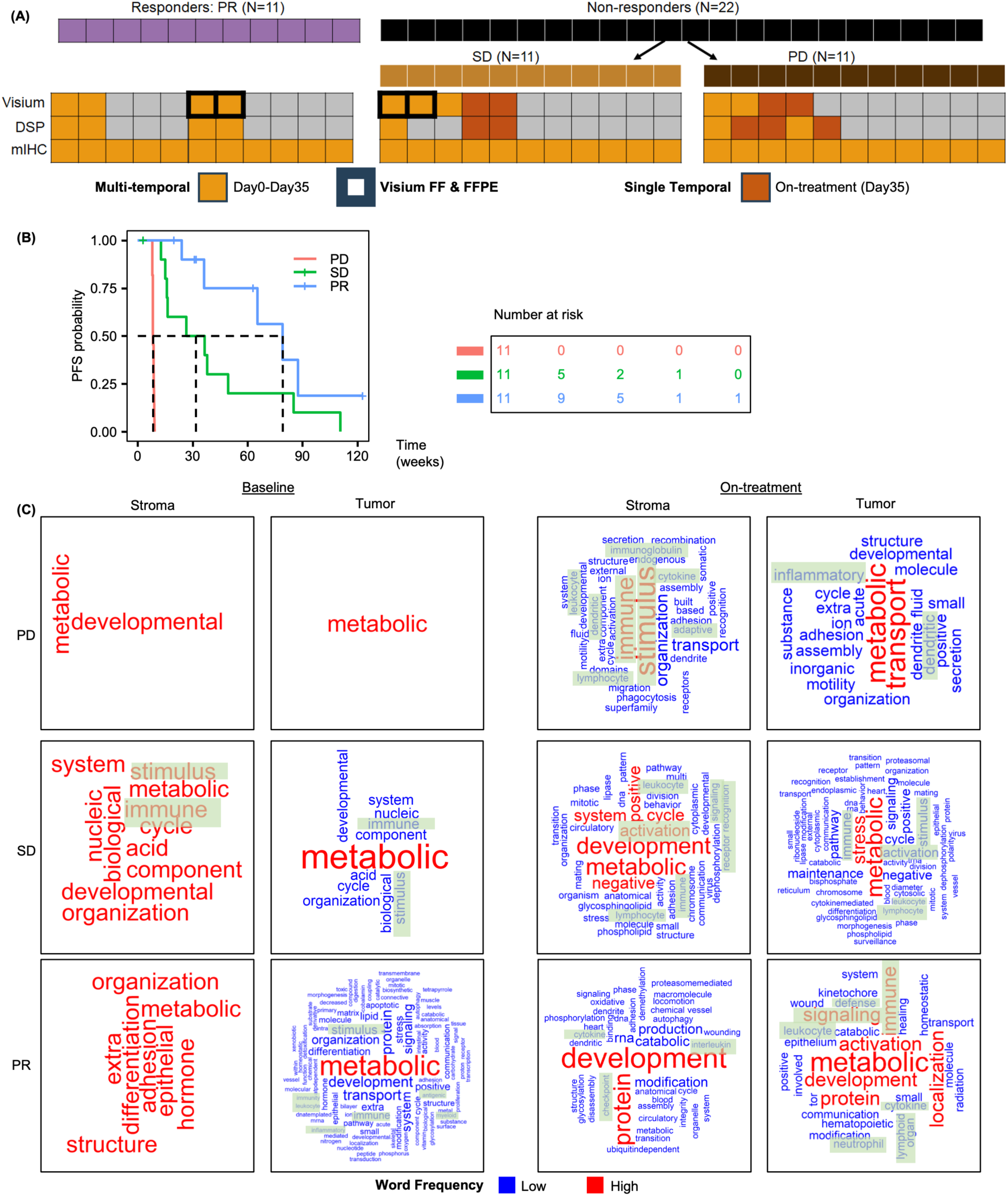
Clinical relevance of differential response groups and spatial omics–based study design. (A) Study design overview, showing spatial omics profiling applied to individual samples collected at multiple time points, grouped by response status. (B) Progression-free survival (PFS) analysis stratified by best response groups: progressive disease (PD), stable disease (SD), and partial response (PR). (C) Tissue region–specific analysis using hdWGCNA, where co-expression gene modules were identified separately in tumor and stroma. Gene modules were subjected to hierarchical GO analysis, with enriched GO terms summarized by response group and visualized as word cloud plots. Terms associated with immune-related processes were highlighted with green boxes.

### Relative Cell Abundance Analysis of mIHC

Relative cell abundance was calculated at the patient level as the proportion of each cell type among all detected cells or CD8⁺ cells (panel details see **Table S1)**. To assess the association between relative cell abundance and clinical response groups (partial response, PR; stable disease, SD; progressive disease, PD), the Kruskal–Wallis test was used to assess overall differences among PR, PD, and SD, followed by post hoc pairwise comparisons (PR vs. PD, PR vs. SD, and SD vs. PD).

### Gene Co-expression Network Analysis

Correlated gene programs in the Visium dataset were identified using, hdWGCNA (v0.4.02). Neighboring spots were aggregated into tissue category–specific metaspots, based on pathologist annotations, using *MetaspotsByGroups*. Optimal soft-thresholding power was determined with *TestSoftPower*, followed by spatial module (SM) construction. Module eigengenes and connectivity were computed using *ModuleEigengenes* and *ModuleConnectivity*, respectively, and visualized using Seurat *DotPlot* and *SpatialFeaturePlot* functions.

### Gene–to-Gene Spatial Analysis in Visium

For PD-1/PD-L1 interactions, Spearman correlation coefficients (*cor*; stats v4.2.2) were computed between spatially adjacent PDCD1 and CD274--expressing spots (raw counts > 1). Immediate neighboring spots were identified using *pairdist* from the spatstat package (v3.0.7), treating spots as a *ppp* object. Correlations were calculated bidirectionally, and the larger coefficient was used to represent PD-1/PD-L1 spatial association. Analyses were performed separately within tumor and stroma.

For POSTN⁺ CAFs and SPP1⁺CD68⁺ macrophage interactions, Visium spots co-expressing POSTN with at least one CAF marker (ACTA2, FAP, PDGFRB, TNC, or S100A4), and spots co-expressing SPP1 and CD68 (raw counts >1), were identified. Spatially adjacent POSTN⁺CAF⁺ and SPP1⁺CD68⁺ spots were identified using *pairdist* from spatstat (v3.0.7). The proportion of interacting neighboring spot pairs was normalized to the total spots within tumor and stroma, respectively. Differences in spatial association metrics were evaluated using the the Kruskal–Wallis test.

### Response-Specific Gene Marker Identification through Low-Dimensional sPLS-DA Modelling

Following principle component analysis (PCA) and partial least squares discriminant analysis (PLS-DA), sparse PLS-DA (sPLS-DA) was performed to identify the most informative gene features, using baseline samples. The tune.splsda function from the mixOmics package was used to optimize feature number, testing gene set sizes from 1-10 and 20-100 (increments of 5). Based on balanced error rate (BER), a two-component model (ncomp = 2) was selected, identifying 105 predictor genes with differential enrichment across response groups.

To identify response-specific gene markers, the 105 sPLS-DA–derived genes were subjected to gene-wise and sample-wise hierarchical clustering. Cluster assignments were determined using *cutree*from the dendextend package (*k = 4*), achieving clear separation of the response groups. Genes uniquely enriched within each response-associated cluster were identified and subjected to Pathway analysis using QIAGEN Ingenuity Pathway Analysis (IPA). To identify a minimal predictive gene set, an outcome-discrimination score was computed as the weighted sum of absolute sPLS-DA gene loadings, weighted by the proportion of response variance explained by each component.

### Data availability

The GEO accession number will be provided upon acceptance. For additional Methods, see Supplementary Information.

## Results

### Discrepancies In Biomarkers Previously Identified By Bulk RNA-Seq

Although bulk omics reveals global molecular changes, they obscure spatial heterogeneity and cellular interactions within the TME. To better characterize localized immune responses to Y90RE-nivolumab, we applied Visium to the same patient cohort from Kaya et al.(9), examining key HCC-related pathways in spatial context. We further stratified the non-responders into PD and SD, as SD demonstrated improved progression-free survival compared to PD (**Fig. 1B**), enabling more refined analysis of patients with no clinical benefit.

We re-evaluated HCC-related pathways and genomic biomarkers. Previously, bulk RNA-seq showed non-responders enriched for Asian-specific Kaya_P2, Hoshida S2, Kaya_M1, Yamashita EpCAM, and Chiang POLYSOMY7 signatures, while responders were enriched in Chiang CTNNB1, Kaya_P1, Kaya_M2, and Hoshida S1 signatures. Now with PD and SD stratification, Chiang CTNNB1, Kaya_M2, and Hoshida S2 were significantly enriched in SD compared to PD, albeit no statistical differences between SD and PR (**Fig. S1A-B**). Amongst genomic biomarkers—similar to prior findings—chromosome 16 deletions increased from PD to PR and *NCOR1* mutation was observed only in PR (**Fig. S1C**).

Visium analysis further revealed discrepancies from bulk RNA-seq at baseline. IFNγ response was reduced in PR compared to PD and SD (**Fig. S2A**), while cell cycle-related pathways (MYC targets and G2M checkpoint) showed similar enrichment across groups. Interestingly, E2F targets were decreased in PR compared to PD and SD but increased in SD relative to PD, indicating distinct profiles between the two non-responder groups that were previously masked when analyzed together. Baseline and on-treatment tissues showed largely consistent pathway enrichment patterns (**Fig. S2B**).

These findings support stratifying non-responders into PD and SD, demonstrating spatial transcriptomics can resolve response-specific signatures obscured in bulk data. These differences prompted further multi-omics analyses to investigate the mechanisms and immune profiles underlying distinct clinical responses.

### Spatial Transcriptomic Reveals Distinct Immune Profiles Across Response Group

Leveraging companion Visium histology images, pathologist-defined tissue annotations were generated for baseline and on-treatment HCC samples from PD, SD, and PR patients. To characterise tissue region-specific spatial transcriptomic (ST) profiles, we performed hdWGCNA to identify GO-annotated gene modules enriched in specific regions and visualized them as word clouds grouped by response category. At baseline, immune responses were detected only in the tumor of PR, while SD showed immune activity throughout the tissue (**Fig. 1C**). In contrast, PD lacked detectable immune response, suggesting intrinsic immune deficiency. On-treatment at day 35, all groups exhibited immune activities in both tumor and stroma, indicating treatment-induced TME modulation. These findings were generally supported by an alternative method, SPATA2(10), despite its reduced pathway resolution (**Fig. S3**).

### Spatial Immune Profiling Reveals T Cell Signals

Following the observed differences in baseline immune activity, we performed quantitative cell-type analysis using enrichment scoring and deconvolution methods in DSP and Visium. We also evaluated spatial niche detection algorithms (stDCL, Proust, and GraphST) using ST data with or without histology images. However, these methods either failed to identify stroma or overestimated tumor areas (**Fig. S4**). Due to these inconsistencies relative to expert annotations, downstream analyses focused on pathologist-defined stromal and tumor regions, representing the most biologically informative compartments for tumor–immune interactions.

At baseline, DSP immune abundance estimation of all-cell regions generally showed low immune cell levels, including T cells, across all response groups (**Fig. 2A**). On-treatment, we expectedly observed increased levels of most immune cells. These findings were similar to those of Visium data (**Fig. 2B**). However, we noted that CD8^+^ T cells remained relatively sparse across tissue regions, as supported by CD8 gene counts. Thus, we applied SPOTlight deconvolution using HCC-specific scRNA-seq–derived cell type reference (11) to characterize T cell subsets with to. At a glance, we observed elevated on-treatment levels of activated CD8⁺ T cells and CD4⁺ memory stem-like cells in SD and PR compared with PD (**Fig. 2C**), suggesting enhanced effective T cell priming and renewal upon treatment. Although baseline CD8⁺ T cell subset levels did not differ significantly across response groups, on-treatment activated CD8⁺ T cell levels were lower in the stroma of PR than SD, but higher in the possibly non-neoplastic region of SD than PD (**Fig. 2D**; both P<0.1). CD4⁺ T cell subsets exhibited greater variation between response group and tissue regions, but CD4⁺ memory stem-like cells were lower in the stroma of SD than PD (P<0.1). Altogether, these observations highlight spatially distinct immune responses across response groups in the on-treatment setting: PR, despite lower CD8^+^ T cell levels than SD, may be characterised by enriched stroma immune cells; SD exhibited CD8^+^ T cell enrichment in non-neoplastic regions, which was not present in PD, suggestive of impaired infiltration and/or retention in PD.

**Fig. 2.**
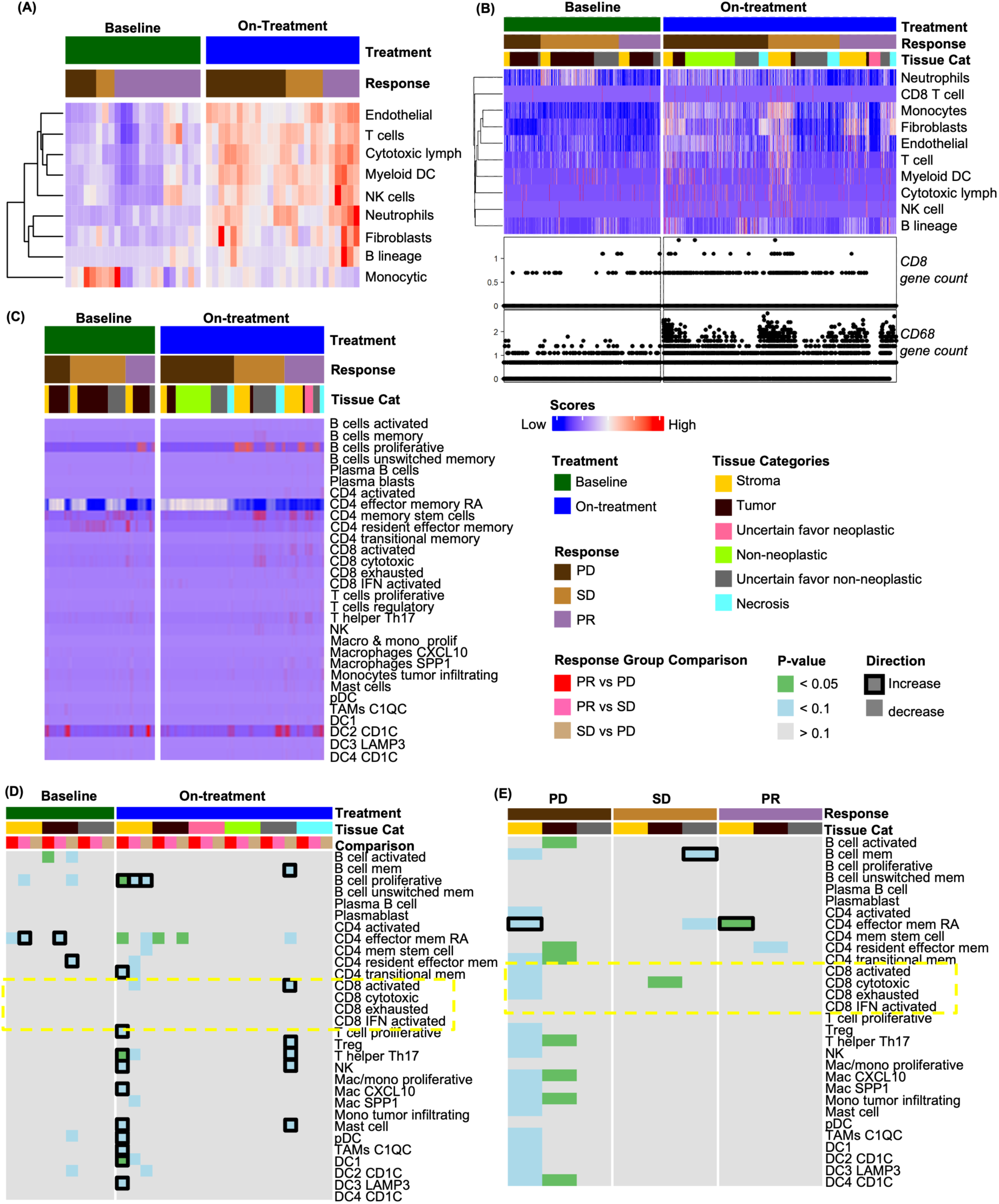
Immune profile characterization at multiple resolutions using spatial transcriptomics. (A) MCPcounter-derived immune cell type scores from DSP data, extracted from ‘General’ (i.e. all-cell) regions, with columns ordered by treatment timepoint and response status; (B) MCPcounter-derived immune cell type scores from Visium data, stratified by tissue category and similarly ordered; (C) Immune deconvolution from Visium data using SPOTlight, based on a detailed single-cell immune reference atlas, with columns ordered by treatment timepoint, response group, and tissue category; (D) Immune abundance differences (from C) across response groups—PD vs SD, PR vs SD, and PR vs PD—assessed using Kruskal-Wallis test; (E) Immune abundance differences (from C) between baseline and on-treatment samples, assessed using Kruskal-Wallis test.

To further delineate treatment effects, we compared differences in immune abundance at baseline and on-treatment, revealing detrimental effects across all response groups, especially in PD (**Fig. 2E**). Significant reduction of immune cells, including activated, cytotoxic, and exhausted CD8⁺ cells, were observed in the tumor (P<0.05) and stroma (P<0.1), potentially reflecting treatment failure to alleviate immune suppression. Although these effects were less pronounced in SD and PR, it is also worth noting that cytotoxic CD8⁺ T cells were decreased (P<0.05) in the tumor of SD, but not in PR, which may partially explain the less durable treatment response. Along with the above findings, we showed that rather than overall immune cell abundance alone, the importance of effective spatial distribution of immune cells in shaping treatment response.

### CD8^+^ T Cell Phenotypes and Spatial Immune Heterogeneity

Persistent antigen stimulation promotes T cell exhaustion, characterised by inhibitory marker expression and impaired effector function. To assess CD8^+^ T cell functional states, we performed single-cell analysis using mIHC to quantify PD-1, LAG-3, and CD38 expression. Although PD-1 is upregulated during early T cell activation, persistent expression represents progression towards T cell exhaustion (12). At baseline, SD exhibited higher PD-1⁺CD8⁺ T cell abundance than PR and PD, suggesting a greater pool of activated T cells that may confer an inherent capacity for treatment response (**Fig. 3A**). On-treatment, despite no significant treatment-associated modulation in the abundance of PD-1⁺CD8⁺ T cells across all response group at the protein level (**Fig. 3B-D**), Visium analysis detected marginal stromal enrichment in SD **Fig. S5A-B**). This could reflect sustained but sub-optimal effector response, resulting in disease stabilisation rather than a durable response.

**Fig. 3.**
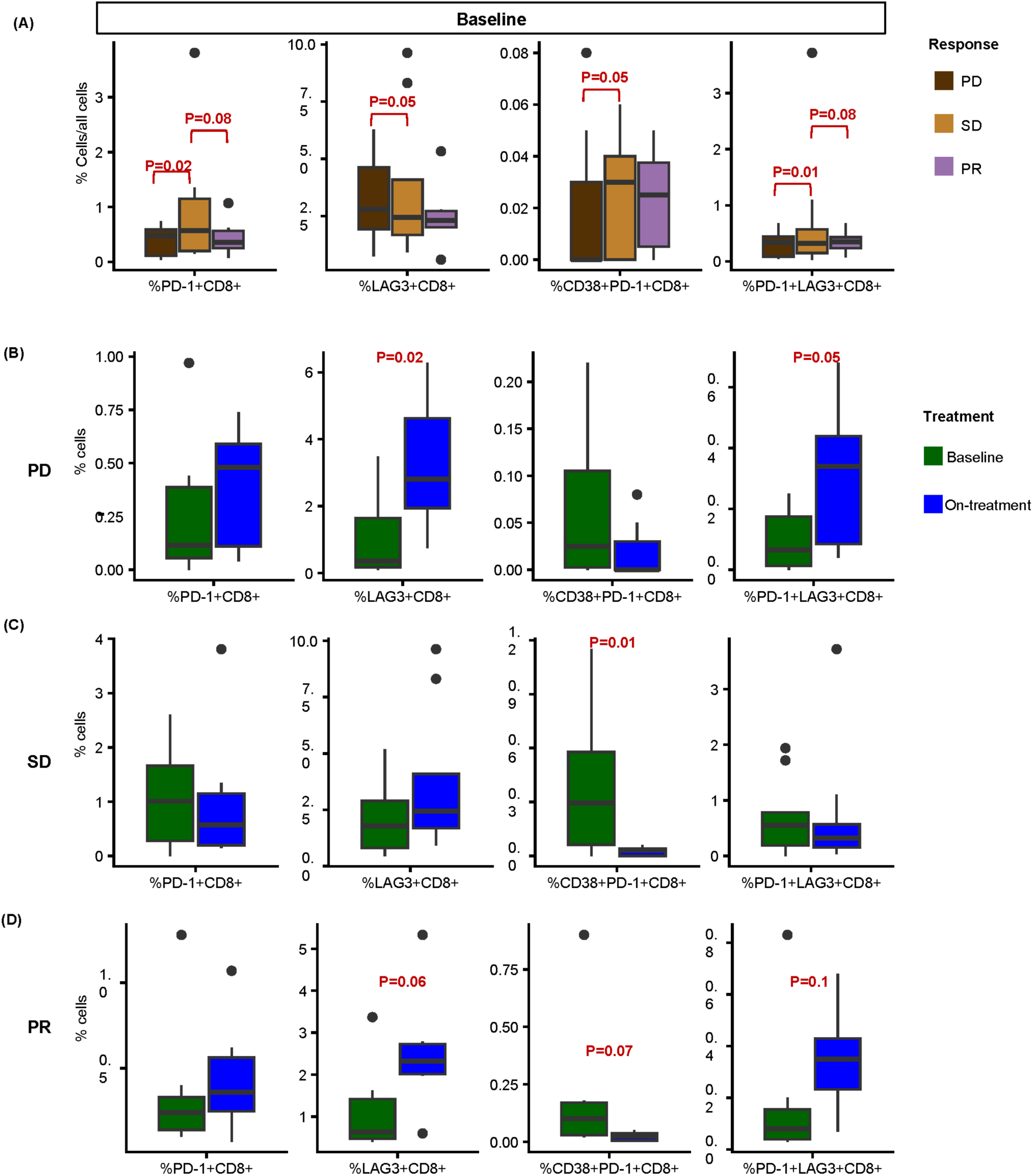
Single-cell analysis of CD8⁺ T cell phenotypes in relation to treatment effect using mIHC. (A) Box plots showing the relative abundance of CD8⁺ subsets (normalized to total cells) across response group comparisons: PR vs. SD, SD vs. PD, and PR vs. PD at baseline. (B–D) Box plots showing the proportions of CD8⁺-related cell types across treatment timepoints within each response group: (B) PD, (C) SD, and (D) PR. Statistical significance was assessed using the Kruskal–Wallis test; p-values > 0.1 are not displayed in panels (A–D).

Further characterising PD-1⁺CD8⁺ T cells, we analysed its co-expression with CD38 and LAG-3. While CD38 is also involved in immune activation, CD38⁺PD-1⁺CD8 T cells represent a subset of exhausted CD8^+^ tissue-resident memory T cells in HCC, often associated with chronic activation and impaired cytotoxic function(13). Baseline percentage of CD38⁺PD-1⁺CD8 T cells were higher in SD than PD (**Fig. 3A**). Treatment-modulated suppression was observed across all response groups, with significant reductions in SD and PR (**Fig. 3B-D**), demonstrating enhanced treatment sensitivity albeit insufficient to reinvigorate effective anti-tumoral response. LAG-3⁺PD-1⁺CD8⁺ T cells, a highly exhausted T cell subset, were more abundant at baseline in SD than PD and PR (**Fig. 3A**), with treatment-modulated enrichment observed in both PD and PR (**Fig. 3B, D**). Conversely, LAG-3⁺CD8⁺ T cells were enriched at baseline in PD but similarly increased in both on-treatment PD and PR. These observations suggest that LAG-3 expression may be dynamically modulated during treatment, including the LAG-3⁺PD-1⁺ population associated with deep exhaustion.

Effective anti-PD-1 therapy depends on spatial interaction and proximity of PD-1 receptors and PD-L1 ligands within the TME. Therefore, we assessed PD-1^/^PD-L1 spatial co-localization using Visium. Baseline levels were similar across stroma and tumor; however, on-treatment stroma PD-1/PD-L1 co-localization was higher in PD than PR (P = 0.06) and marginally lower in SD than PR (P = 0.14) (**Fig. S5C**). Despite a trend of increased and decreased correlation in the stroma of PD and SD, respectively, there was no significant treatment-modulated effects on PD-1/PD-L1 co-localization within individual response group (**Fig. S5D**). This finding in PD, when coupled with the absence of a significant PD-1⁺CD8⁺ T cell population at baseline, suggests a lack of functional targets for PD-1 blockade—potentially contributing to diminished responsiveness to anti–PD-1 therapy. This may reflect immune evasion through sparse PD-1⁺ T cell infiltration and could be partially driven by an immunosuppressive microenvironment involving alternative inhibitory pathways such as LAG-3 (**Fig. 3B**), indicating a treatment-induced shift toward compensatory exhaustion mechanisms that could be alleviated through combination strategies with anti–LAG-3 agents(14).

Beyond biomarker findings, we further performed cell-type-specific differential expression analysis using DSP on CD8^+^ AOIs to investigate on the functional state of CD8^+^ T cells. In SD compared to PD, CD8⁺ T cells had upregulated antigen presentation via MHC and αβ T cell receptors, adaptive immune responses, activation, and co-stimulatory signaling pathways (**Fig. S6A**). No gene ontology terms associated with negative regulation of T cells were detected, suggesting that the elevated PD-1⁺CD8⁺ T cells in SD were unlikely to represent exhausted cells, but rather reflect an activated state, albeit sub-optimum(15). On-treatment, there were no significant differences in immune regulation across response groups (**Fig. S6B**), consistent with earlier Visium findings (**Fig. 2D**). These findings suggest that SD reflects partial immune activation, in which PD-1⁺CD8⁺ T cells are present and treatment responsive but with impaired intratumoral infiltration, therefore limiting anti-tumor efficacy.

### Macrophage Phenotypes and Spatial Immune Heterogeneity

The macrophage axis represent an additional potentially important driver of therapeutic response(9). DSP analysis first revealed higher levels of monocytic cells at baseline, with a notable reduction on-treatment, in PD and SD. (**Fig. 2A**). Visium analysis, however, showed on-treatment increase in monocytes and robust CD68 gene count in the stroma and tumor (**Fig. 2B**). This discrepancy may reflect differences in spatial sampling, where DSP signals may have been diluted when averaged across broader tissue areas or tumor-enriched regions. SPP1+ macrophages, which were implicated in forming the immunosuppressive tumor immune barrier (TIB)(16), had enriched baseline levels in the stroma of PD and SD as compared to PR, but similar levels in the tumor of all response group (**Fig. 4A**). On-treatment, percentage of SPP1⁺ macrophages was marginally lower in the tumor of PR than SD (P=0.14) and slight treatment-modulated reduction was observed within the tumor of PR (P=0.15, **Fig. 4B**), hinting at sub-optimal treatment responsiveness. However, we were unable to demonstrate the localization of SPP1⁺ cells at the invasive margin and the physical structure of the TIB, as reported in previous studies, in our baseline and on-treatment tissues from PD and SD (**Fig. 4C-D**). Instead, SPP1⁺ macrophages were broadly distributed across both stroma and tumor. We postulate that this observation is partly due to the nature of the biopsy-derived tissue samples, which lacks an invasive margin.

**Fig. 4.**
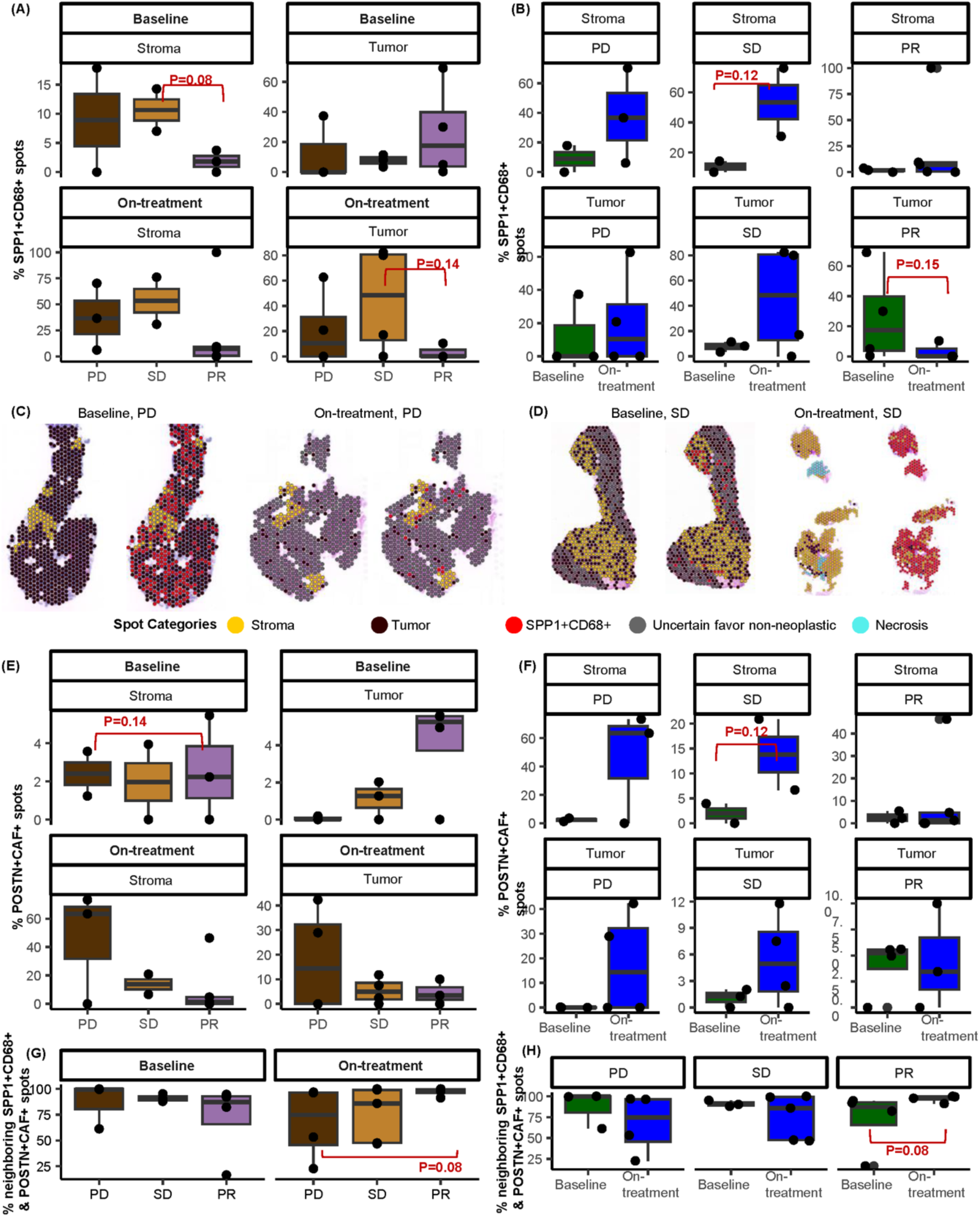
Spatial analysis of SPP1⁺CD68⁺ macrophages and POSTN⁺CAF⁺ fibroblasts stratified by tissue category using Visium. (A–B) Comparison of the percentage of SPP1⁺CD68⁺ spots across response groups: (A) PR vs SD, SD vs PD, and PR vs PD; (B) baseline vs on-treatment.(C–D) Representative tissue maps showing the localization of SPP1⁺CD68⁺ spots in tumor versus stroma from paired pre- and post-treatment samples of a (C) PD and (D) SD case, paired pre-post samples. (E–F) Comparison of the percentage of POSTN⁺CAF⁺ spots across response groups: (E) PR vs SD, SD vs PD, and PR vs PD; (F) baseline vs on-treatment. (G–H) Spatial correlation between SPP1⁺CD68⁺ macrophages and POSTN⁺CAF⁺ populations, measured as the percentage of spatially co-localizing spots relative to the total number of Visium spots: (G) across response groups—PR vs SD, SD vs PD, and PR vs PD; (H) baseline vs on-treatment.

Applying the same analyses to POSTN⁺ cancer-associated fibroblasts (CAFs), a population proposed to recruit macrophages and induce SPP1 expression(17), we showed that baseline percentages were similar in the stroma across all response groups, although it was marginally higher in the tumor of a subset of PR (**Fig. 4E**). Treatment did not significantly impact POSTN⁺ CAFs (**Fig. 4F**). Additional analysis further revealed that treatment increased spatial correlation of POSTN⁺ CAFs and SPP1⁺ macrophages in PR (**Fig. 4G–H**), thereby limiting the above notion on treatment efficacy and potential disruption of the TIB. Together, these findings suggests that the SPP1 macrophage and POSTN CAFs axis alone does not account for treatment response and is unlikely the primary to be the primary determinant of therapeutic efficacy. To further investigate this, we analysed the abundance and spatial distribution of other CD68⁺ macrophage subtypes.

Protein level mIHC analysis revealed significantly higher baseline percentages of CD38⁺CD68^+^, CXCL9⁺CD68^+^, and CXCR3⁺CD68^+^ macrophages in PR compared PD and SD (**Fig. 5A**). Additionally, CXCL9⁺CD68^+^ and CXCR3⁺CD68^+^ macrophages, but not CD38⁺CD68^+^ macrophages, were observed in close proximity to tumor cells within PR (**Fig. 5B-E**), suggesting closer involvement in anti-tumor immunity. This spatial distribution suggests that durable response shown in PR may be driven by the selective intratumoral localization of certain macrophages. On-treatment, overall percentage of CD68^+^ macrophages were higher in PD than PR, and CD38⁺CD68^+^ macrophages were significantly more abundant in PD than SD (**Fig. 5F**). Treatment-induced effect differed by response status; PD had increased CD38^+^CD68^+^ and CXCL9^+^CD68^+^ macrophages but reduced CXCR3^+^CD68^+^ macrophages, whereas SD showed selective increase in CXCL9^+^CD68^+^ macrophages only, and PR exhibited reduced CXCR3^+^CD68^+^ macrophages only (**Fig. 5G**). Taken together, these findings suggest that CD38^+^CD68^+^ macrophages are associated with treatment resistance, whereas CXCL9^+^CD68^+^ and CXCR3^+^CD68^+^ macrophages are potentially treatment responsive but unable to independently sustain durable therapeutic benefit.

**Fig. 5.**
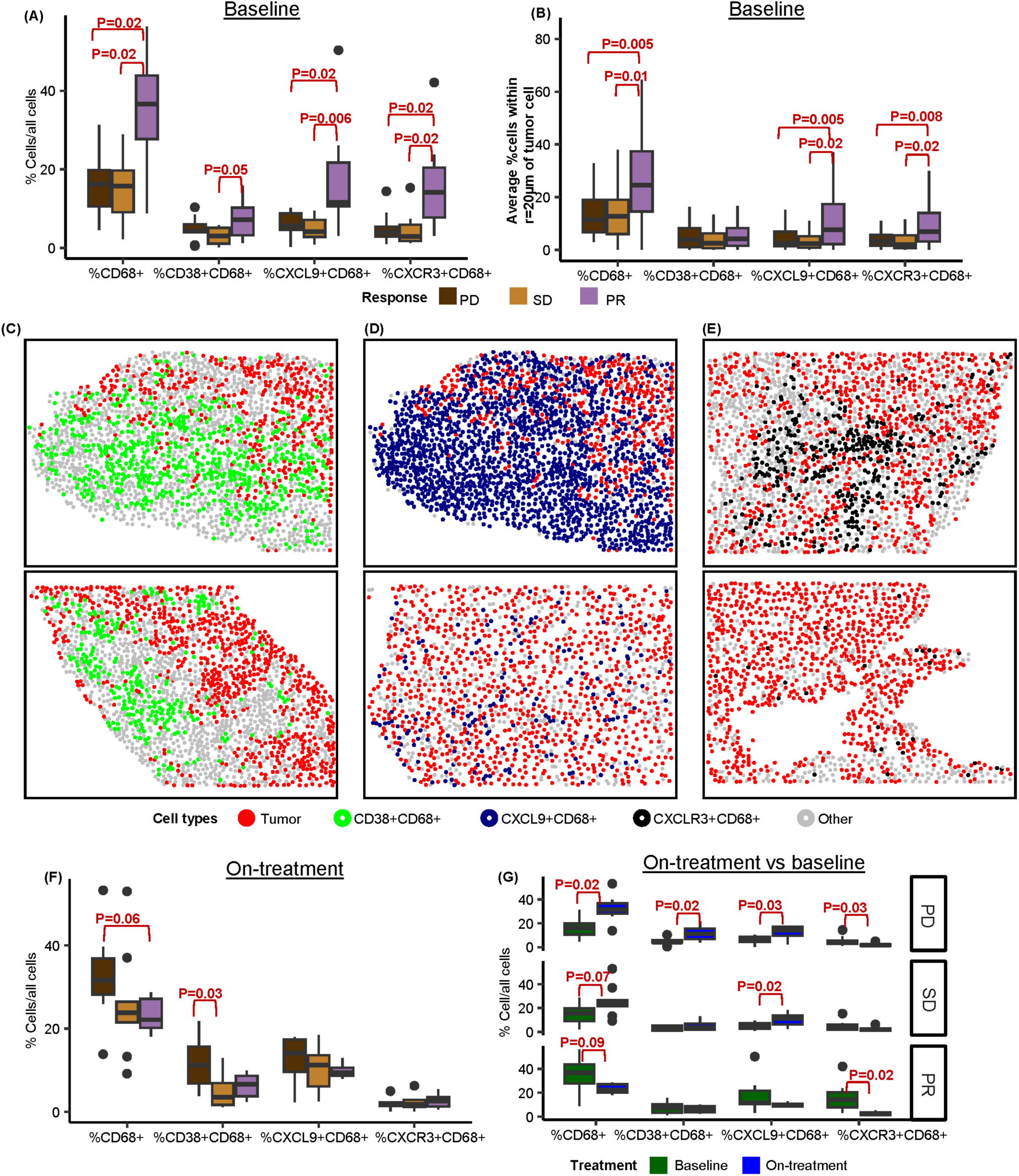
Relative abundance and spatial proximity of CD68⁺ phenotypes to tumor cells, assessed using mIHC. (A) Comparison of the relative abundance of various CD68⁺ phenotypes across response groups (PD, SD, PR) at baseline. (B) Proportion of CD68⁺ phenotypes located within 20 µm of tumor cells across response groups at baseline. (C–E) Representative tissue maps showing the spatial distribution of (C) CD38⁺CD68⁺, (D) CXCL9⁺CD68⁺ , and (E) CXCR3⁺CD68⁺ cells in relation to tumor from selected PR cases. The top panel shows high-density tissue samples, while the bottom panel shows low-density samples. (F) On-treatment abundance differences and (G) treatment-associated changes in CD68⁺ phenotypes across response groups. Linear mixed-effects model was used for statistical analysis in (A), (B), (F), and (G).

With the increased abundance of tumor-proximal CD68⁺ macrophages in PR versus PD and SD (**Fig. 5A–B**), we performed DSP on CD68^+^ AOIs to investigate on its functional state, beyond cell abundance. Interestingly, baseline CD68^+^ cells in PR were enriched in humoral immune pathways, suggesting the possibility of macrophage-B-cell crosstalk (**Fig. S6C**). This is consistent with pan-cancer observations linking high CD68 expression to B cell abundance(18), although there was no additional treatment-associated immune modulation observed in PR. Instead, CD68⁺ macrophages in SD had upregulated cytokine production, enhanced humoral immune response and increased leukocyte-mediated activity than in PD (**Fig. S6D**).

Collectively, we showed that the TME of PD is characterized by the enrichment of CD38^+^CD68^+^ and CXCL9^+^CD68^+^ macrophages that are non-responsive to treatment. CXCL9^+^CD68^+^ macrophages were functionally enriched in SD, but CXCR3^+^CD68^+^ macrophages were reduced in PR. These observations suggests an immunosuppressive role of CD38^+^CD68^+^ macrophages, and hint at the role of the CXCL9-CXCR3 axis, which is known to regulate immune reactivity.

### Transcriptomic Signature Predicts Treatment Unresponsiveness

Given the clinical importance of predicting treatment response, we next sought to identify predictive signatures capable of stratifying PD from SD, using baseline samples. This may enable early identification of patients unlikely to benefit from therapy, thereby avoiding unnecessary therapy and informing alternative combination approaches. PCA and PLS analyses demonstrated that DSP relative to Visium, provided the appropriate spatial resolution for segregating transcriptomic profiles across response groups (see Supplementary Information). We therefore applied sPLS-DA to identify the most discriminative genes, revealing 72, 19, and 14 genes associated with PD, SD, and PR, respectively (**Table S2**). Gene expression patterns in PD and SD were relatively homogeneous and pronounced, but PR-associated genes exhibited greater heterogeneity and lower expression levels (**Fig. 6A**). Pathway analysis of the 72 baseline PD-associated genes revealed enrichment of stress-adaptation pathways (EIF2AK4/GCN2, EIF2, ATF4, NMD), growth factor-driven resistance pathways (mTOR, MAPK–PI3K), angiogenic/fibrotic remodeling pathways (EMT, VEGF/HIF1α), and immune exclusion/exhaustion pathways (IFNα/β, IL-10, WNT/β-catenin) (**Table S3**).

**Fig. 6.**
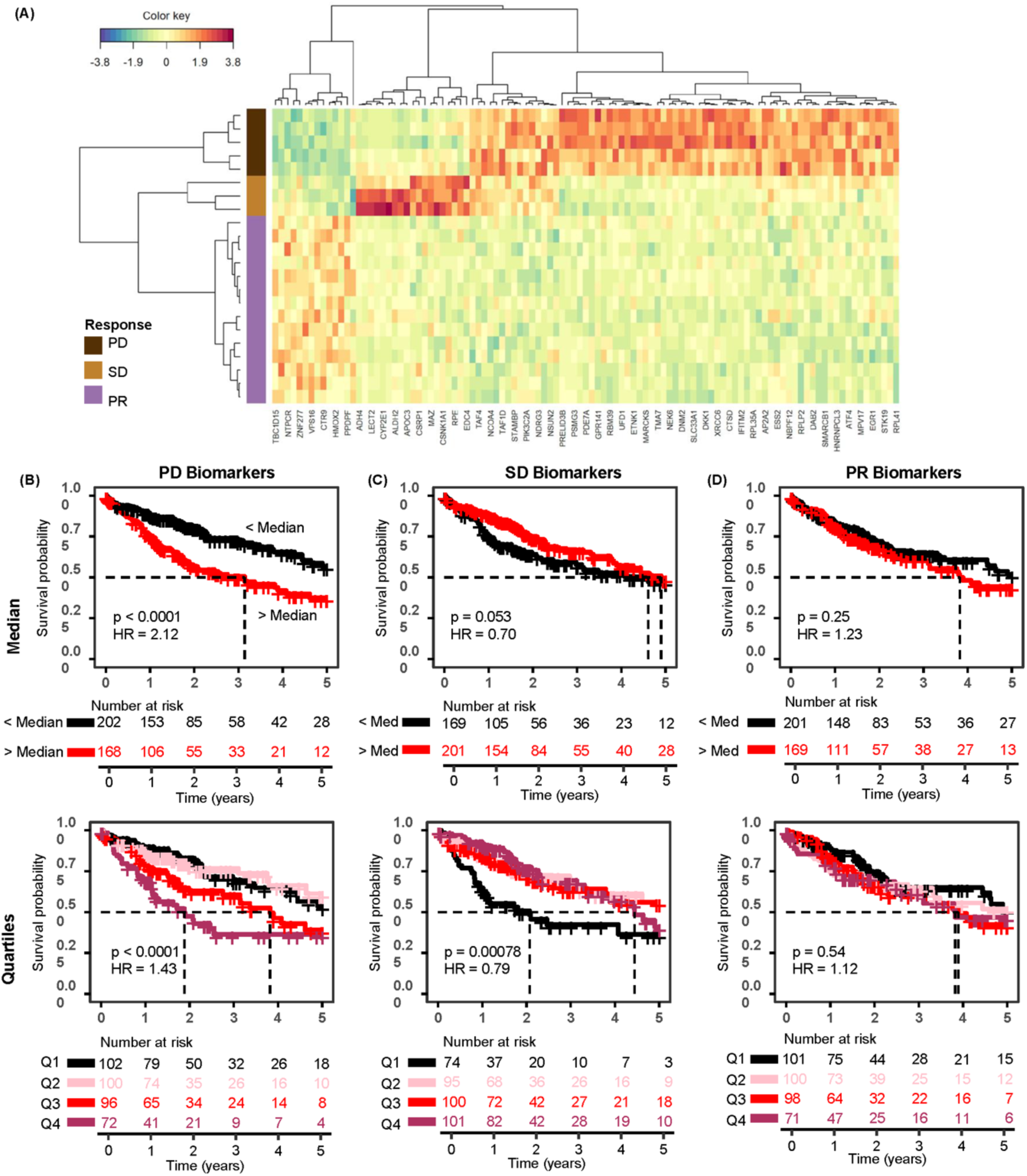
Response-specific transcriptomic profiles identified by sPLS-DA using DSP data. (A) Heatmap showing differential expression of response-specific genes, with rows representing response groups. (B–D) Stratification of patients from the TCGA LIHC cohort based on enrichment scores of genes upregulated in (B) PD, (C) SD, and (D) PR groups. Stratification was performed using either median (B-D: top) or quartile thresholds (B-D: bottom). Kaplan–Meier survival analysis and log-rank test were used to assess survival associations among these groups.

We further evaluated the prognostic relevance of these gene signatures in the public TCGA Liver Hepatocellular Carcinoma (LIHC) cohort. Kaplan-Meier analysis, using both median and quartile stratification, revealed that higher PD signature expression was associated with significantly poorer survival (both P<0.0001; **Fig. 6B**) while higher SD signature expression correlated with favourable prognosis (P=0.053, P=0.00078; **Fig. 6C**). In contrast, the PR signature showed no significant prognostic association (P=0.25, P=0.54; **Fig. 6D**). Using the outcome-discrimination scoring approach, the top three most discriminative PD signature genes (*DNM2*, *SLC33A1*, and *RPL37*) retained prognostic significance in the TCGA cohort, although with attenuated association (P=0.016). Although treatment information was unavailable in TCGA-LIHC, these findings support the clinical relevance of the PD signature. The inherently immune-deficient TME associated with poor response to Y90RE–Nivolumab in PD may reflect an intrinsically resistant state linked to broader unfavourable outcomes, underscoring the need for additional therapeutic targeting baseline immunosuppression.

## Discussion

The combination of Y90RE and ICI shows promise in a subset of patients with advanced HCC; however, early identification of patients with no clinical benefit, particularly those with PD, remains a critical unmet need. To address the limited understanding of the TME and lack of predictive biomarkers, we applied an integrated multi-omics framework combining ST (Visium and DSP), histology-based annotations, and proteomic profiling (mIHC) to analyze a Y90RE–nivolumab–treated HCC cohort.

Treatment resistance in PD appears to be driven by impaired CD8^+^ T cell infiltration and functional suppression, coupled with functionally suppressed CD68^+^ macrophages, that collectively sustains an immunosuppressive TME. We showed that baseline CD8^+^ T cells had diminished capacity to elicit adaptive immune responses and co-stimulatory signaling, accompanied by the lack of on-treatment CD8^+^ T cell enrichment and a significant reduction of various immune cells, including activated, cytotoxic, and exhausted CD8^+^ T cells. Baseline phenotyping further revealed low levels of PD-1^+^CD8^+^ and LAG-3⁺PD-1⁺CD8⁺ T cells but high level of LAG-3^+^CD8^+^ T cells, suggesting an inherently reduced pool of functional target for Y90RE-Nivolumab. The enrichment of both LAG-3-expressing subsets on-treatment further suggests a compensatory reliance on alternative checkpoint pathway. Although prior studies in advanced HCC treated with only ICI, have associated high baseline levels of LAG-3^+^ and LAG-3^+^CD8^+^ T cells with improved prognosis and response to PD-1 blockade(19), our findings showcase a treatment-induced compensatory shift to LAG-3 signalling, which may underlie treatment resistance. We hypothesize that the discrepancy was due to co-expression with PD-1, as double positive LAG-3^+^PD-1^+^CD8^+^ T cells likely reflect deeper exhaustion states(20, 21) and limited rescue potential.

Supporting this notion, studies have demonstrated synergistic activity of LAG-3 and PD-1 co-expression in driving CD8^+^ T cell exhaustion via upregulation of TOX, a transcription factor that establishes terminal exhaustion(21, 22). Since the TME in PD is characterised by terminal exhaustion and alternative checkpoint dominance, along with FDA approval of combined nivolumab and relatlimab treatment for melanoma(23), our findings provide biological rationale for exploring dual PD-1 and LAG-3 blockade in advanced HCC enriched for LAG3⁺PD-1⁺ T cells, an approach currently being investigated in a recently completed RELATIVITY-073 trial (NCT04567615), with results pending(24).

Macrophage dysfunction may also reinforce therapeutic resistance. While PD exhibited the lowest baseline abundance of CD68^+^ macrophages, Y90RE-Nivolumab selectively induced expansion of CD38^+^CD68^+^ and CXCL9^+^CD68^+^ macrophages. Notably, CD68⁺ macrophages in PD appeared functionally attenuated, exhibiting reduced cytokine production, diminished humoral responses, and lower leukocyte-mediated activity compared with SD. This contrasts with previous reports associating CD38^+^CD68^+^ macrophages with anti-tumoral activity, favourable prognosis following surgical resection in early-stage HCC(25), and improved response and survival following ICI treatment in advanced HCC(26). These observations suggest that macrophage functional phenotypes may be influenced by disease stage, treatment modality, and/or TME-mediated suppression. notably, CD38 also function as an ectoenzyme that initiates the CD38/CD203a/CD73 adenosinergic pathway, generating adenosine that suppresses T cells, natural killer cells, and dendritic cells(27, 28). Therefore, in advanced HCC treated with Y90RE-Nivolumab, CD38 expression may reflect metabolic dysfunction and immunosuppression rather than simple immune activation.

Extending previous bulk analyses that identified higher abundance of CXCL9⁺CD68⁺ and CXCR3⁺CD68⁺ macrophages in responders as potential biomarkers(9), our spatial analyses further revealed elevated baseline levels of tumor-proximal CXCL9⁺CD68⁺ and CXCR3⁺CD68⁺ macrophages in PR compared with PD and SD. We further observed distinct on-treatment remodelling suggestive of disrupted CXCL9-CXCR3 signalling across response groups. Although both PD and SD were enriched for CXCL9^+^CD68^+^ macrophages, PD showed reduced CXCR3^+^CD68^+^ macrophages, whereas SD macrophages exhibited upregulated cytokine production, leukocyte-mediated activity, and humoral immune response. In contrast, PR displayed selective suppression of CXCR3^+^CD68^+^ macrophages. Considering the role of CXCL9-CXCR3 axis in regulating immune reactivity(29), these findings highlight its importance in shaping anti-tumor immunity and potentially treatment responsiveness.

Our study highlighted LAG-3-mediated T cell exhaustion and macrophage dysfunction as immune features potentially underlying treatment resistance. Together with the modest 30% objective response rate previously reported for this cohort(5), we identified and validated a 72-gene PD signature suggesting that baseline PD tumors are primed toward tumor-intrinsic stress adaptation, angiogenic/fibrotic remodeling, and immune exclusion/exhaustion. Enrichment of EIF2AK4/GCN2, EIF2, ATF4, NMD, MAPK–PI3K/mTOR, EMT, and VEGF/HIF1α pathways are suggestive of a hypoxic, integrated stress response(30), growth factor–driven, and stroma/angiogenic phenotype that may impair CD8⁺ T cell infiltration or activation(31), consistent with our findings of low PD1⁺CD8⁺ levels. Concurrent enrichment of IFNα/β, IL-10, WNT/β-catenin, and co-inhibitory pathways suggests compensatory immune suppression and exhausted CD8⁺ states(32), consistent with elevated levels of LAG3⁺CD8⁺ cells. Notably, WNT/β-catenin signaling is a well-established immune exclusion mechanism linked to poor response to PD-1/PD-L1 blockade. Among the three most discriminative PD-associated genes, ribosomal protein L37 (*RPL37*) was consistently co-enriched with stress adaptation and translational control pathways, including EIF2AK4/GCN2, EIF2, ATF4, and NMD(30). High *RPL37* expression associated with worse survival in HCC(33), and increased expression in Regorafenib-resistant colorectal cancer cells suggest a role in mediating acquired resistance(34). Solute carrier family 33 member 1 (*SLC33A1*) promotes tumor expansion through GCN2-mediated integrated stress responses and regulation of amino acid transporter genes(30), and is highly overexpressed in HCC(35). Dynamin 2 (*DNM2*), albeit not enriched in significant pathways, was reported to facilitate LAMTOR1-mediated endocytosis of surface MHC-II molecules, impairing CD4⁺ T cell antigen presentation and downstream anti-tumoral CD8⁺ T cell activation (36). This represents a key mechanism to immune evasion and acquired resistance to immunotherapeutic interventions(37). Collectively, upon further validation, this gene signature could serve as a potential tool for improving patient stratification and outcome prediction.

Several limitations of this study should be acknowledged. First, the multi-cell resolution and relatively low read depth of Visium limited deep immune phenotyping. To mitigate this, we complemented Visium with orthogonal spatial modalities, including single-cell–level phenotyping by mIHC and cell type–resolved functional transcriptomic profiling by DSP, providing higher resolution characterization of immune cell states and functions. Second, the analysis of the SPP1⁺ macrophage barrier and its spatial association with POSTN⁺ CAFs should be considered exploratory, as biopsy samples lacked clearly defined tumor margins. This highlights a broader challenge of barrier-based spatial analyses in biopsy specimens, where resection specimens preserving full tissue architecture are often unavailable. Third, the small cohort size precluded stratified analyses by etiological factors such as viral status, which may influence immune responses and treatment outcomes in HCC. Future studies should validate the 72-gene PD-associated signature in independent ICI-treated cohorts to determine whether it reflects a pre-existing immune-resistant TME linked to unfavourable responses. Such validation may enable early identification of patients unlikely to benefit and inform alternative therapeutic strategies, including combination approaches targeting additional immune checkpoints such as LAG-3.

## Data Availability

The GEO accession number will be provided upon manuscript acceptance.

## Acknowledgments

We acknowledge Neslihan Arife Kaya for providing bulk omics data and Laurens Lehner for contributions to data analysis. The results shown here are in part based upon data generated by the TCGA Research Network: https://www.cancer.gov/tcga.

## Financial support and sponsorship

This work was supported by the BII and SIgN (A*STAR) core fund to M.C.L.; by National Medical Research Council the Open Fund–Young Individual Research Grant (MOH-OFYIRG23jan-0049 to M.C.L.); by A*STAR Council Strategic Fund BMRC (BII_C250314072 to M.C.L.); by National Medical Research Council (OFLCG23may-0039, OFLCG24MAY-0028, OFLCG24may-0025, CIAINV25jan-0001, MOH-001067, MOH-001610-00, OFLCG23may-0031); and by A*STAR GAP Funding (I24D1AG059).

## Conflict of interest

nothing to report.

## Abbreviations

BER: balanced error rate
CAFs: cancer-associated fibroblasts
DSP: Digital Spatial Profiler
HCC: hepatocellular carcinoma
ICI: immune checkpoint inhibitors
IPA: Ingenuity Pathway Analysis
LIHC: Liver Hepatocellular Carcinoma
mIHC: multiplex immunohistochemistry
PCA: principle component analysis
PD: progressive disease
PLS-DA: partial least squares discriminant analysis
PR: partial response
SD: stable disease
SM: spatial module
sPLS-DA: sparse PLS-DA
ST: spatial transcriptomic
TME: tumor microenvironment
Y90RE-Nivolumab: Y90RE with Nivolumab
Y90RE: yttrium-90.

## Author contributions

D.T. and J.Y. conceived and designed the study. M.C.L., M.Z., M.P.R., Z.Y.C., and X.Y.W. contributed to data analysis and interpretation. Z.W.N. and X.L. collected tumor tissue samples and performed the omics experiments. J.P.V. and J.Y. conducted histopathological analysis and provided annotations. M.C.L. and D.G. drafted the manuscript. D.T. and J.Y. contributed to editing and critical revision of the manuscript for important intellectual content. All authors reviewed and approved the final manuscript.

## Notes

### Competing Interest Statement

The authors have declared no competing interest.

### Author Declarations

IRB of the Agency for Science, Technology and Research (A*STAR), (IRB: 2021-112) gave ethical approval for this work.

