## Supplementary Information for "Spatial Analysis Uncovers Immune Resistance Mechanisms in Non-Beneficial Hepatocellular Carcinoma Treated with Y90 Radioembolization-Nivolumab"

Patient Cohort

The clinical trial (CA 209–678) was a single-arm, single-center, two-stage phase 2 trial designed to investigate the activity and safety of Y90-radioembolization followed by nivolumab in patients with advanced HCC. Participants provided written informed consent [1]. Thirty-six patients with Child-Pugh A cirrhosis and advanced HCC not suitable for curative surgery were recruited from the National Cancer Centre Singapore/Singapore General Hospital, Singapore. The full inclusion/exclusion criteria and response determination based on the RECIST method can be found in the original study report [1].

For each of the 33 patients with available treatment response data (out of 36 enrolled in the trial), both pretreatment/baseline and on-treatment biopsies were collected from the same Y90-exposed liver lesion before Y90-RE (Day 0) and after one dose of intravenous nivolumab (Day 35) [2]. All these 33 baseline and on-treatment tissue samples went sent for multiplexed immunohistochemistry (mIHC); a subset of 13 patient samples were sent for 10x Visium profiling; a subset of 12 patient samples were sent for DSP profiling (Fig.1A).

Progression-Free Survival Analysis by Best Overall Response

Best overall response (BOR) was defined as the best response recorded from the start of the treatment until disease progression/recurrence using criteria of RECIST 1.1 (i.e., Complete Response (CR), Partial Response (PR), Stable Disease (SD), Progressive Disease (PD)). The patient's best response assignment depended on the achievement of both measurement and confirmation scan.

Time to progression (TTP) was defined as time from study entry until the first date that progressive disease was objectively documented. Patients with no documentation of progressive disease were censored at the date of last tumour assessment.

Progression-free survival (PFS) was defined as time from study entry until objective tumour progression, or death from any cause, whichever occurs first. Patients who were alive or did not have an assessment of PD were censored at the date of last tumour assessment.

The survival distributions were estimated and compared among patients with different BOR categories (PD, SD, and PR) using the Kaplan–Meier method. The analyses were conducted using the survfit function from the survival package in R 4.4.3.

Histology Quality Check (QC) Prior To 10x Visium Profiling

Formalin-Fixed Paraffin-Embedded (FFPE) tissues samples were sectioned at thickness of 4 um and stained with the in-house clinically validated Hematoxylin and Eosin stain (Epredia, Kalamazoo, MI, USA). The stained sections were imaged at 200x magnification using Zeiss AxioScan 7 with its corresponding Zeiss Zen software (version 3.7; Carl Zeiss Microscopy, Jena, Germany) and the images were assessed for their sample morphology and the presence of the Y90 beads.

10x Visium Profiling

Visium Spatial Gene Expression (10x Genomics, Pleasanton, CA, USA) relies on the use of specialized Visium slides that consist of four capture areas which contain about 5,000 barcoded spots each. These spots in turn contain many oligonucleotides, resulting in millions per capture area. Each oligonucleotide consists of a 30-nucleotide poly(dT) sequence for the capture of polyadenylated mRNA molecules, a 12-nucleotide unique molecular identifier (UMI), a 16-nucleotide spatial barcode (a unique set of spatial barcodes is shared by all oligonucleotides within each individual gene expression spot, abbreviated as a Visium spot), and a partial TruSeq Read 1 sequence for library preparation and sequencing.

FFPE sectioned tissues (5-µm thick) were placed on the capture areas (each 6.5 mm × 6.5 mm) of the Visium Gene Expression slide (10x Genomics, Pleasanton, CA, USA), heat-fixed at 42 C on a thermocycler and air-dried overnight in a dessicator. The slides were then stained with Mayer’s Hematoxylin (Agilent, Santa Clara, CA, USA) followed by Eosin Y solution (Abcam, Cambridge, UK) and imaged at 200x magnification using Zeiss AxioScan 7 and Zeiss Zen software (version 3.7; Carl Zeiss Microscopy, Jena, Germany). Image conversion to TIFF format at 10% size was done to facilitate Visium data processing. Using the Visium FFPE Reagents Kit and Visium Human Transcriptome Probe Kit (10x Genomics, Pleasanton, CA, USA), the tissue sections then underwent mRNA probe hybridization, ligation and permeabilization. The released mRNA molecules were then captured by the poly(dT) sequence on the Visium slide surface and reverse transcribed, leaving cDNA molecules complementary to the spatial barcode and UMI covalently attached to the slide. The spatially barcoded molecules were released from the slide, underwent library preparation, purification and PCR-amplification using the Visium Library Preparation Kit and Dual Index Kit TS Set A (10x Genomics, Pleasanton, CA, USA). The purified cDNA libraries were then sequenced at the recommended depth of 50 K raw reads per capture spot covered by the tissue.

High-Resolution H&E Image Acquisition for Pathologist Annotation

The Zeiss AxioScan 7 imaging system and its pairing Zeiss Zen software (version 3.7; Carl Zeiss Microscopy, Jena, Germany) were used for the image acquisition of the hematoxylin and eosin-stained samples at 200x magnification, producing high-resolution images suitable for the pathologist annotations. Images were subsequently converted to TIFF format at 10% of the original size for Visium data processing.

NanoString GeoMx Digital Spatial Profiling (DSP) – Transcriptomics

NanoString spatial profiling [3, 4] was used to analyse 3 protein-level and 1,800 transcriptomic-level immune-markers simultaneously on FFPE tissue slides. Following deparaffinization and antigen retrieval procedures, sections were simultaneously incubated overnight with fluorescent-labeled antibodies against CD8 and CD68.

After staining, the tissues were scanned using a GeoMx DSP instrument to generate high-resolution digital fluorescent images. Within each slide, multiple regions of interest (ROIs) were selected by pathologist (JY). Each ROI was either collected as a whole region or segmented into Areas of Illumination (AOIs) for compartment-specific profiling. AOI segmentation was marker-guided, whereby CD8⁺ and CD68⁺ AOIs were defined based on immunofluorescence signals and collected independently.

For oligonucleotide collection, UV light was directed through a programmable digital micromirror device (DMD) or dual DMD (DDMD) to precisely illuminate the ROI or AOI and cleave the photocleavable oligos (PC-oligos). Released oligos were then collected by microcapillary tube inspiration and dispensed into a 96-well plate. Digital quantification of the collected oligos was performed using the nCounter System, enabling single-molecule counting [5][6].

mIHC Staining, Image Quality Check and Preprocessing

mIHC staining was performed on 4-µm thick FFPE tissue sections using Leica Bond Max autostainer (Leica Biosystems, Melbourne, Australia), Bond Refine Detection Kit (Leica Biosystems, Newcastle Upon Tyne, UK) and Opal Fluorophore Reagent Packs (Akoya Biosciences, Marlborough, MA, USA)[7-10].

In brief, FFPE tissue sections were deparaffinised, rehydrated and thereafter, subjected to repeated cycles of heat-induced epitope retrieval, incubation with primary antibody, secondary antibodies conjugated with polymeric HRP (Bond Refine Kit) and Opal tyramide signal amplification (TSA) (Akoya Biosciences, Marlborough, MA, USA). Two separate 7-plex panels were profiled: Panel 1: DAPI, CD8, LAG3, CD38, PD-1, PD-L1, STAT1; Panel 2: DAPI, CD45, CD8, CD68, CD38, CXCR3, CXCL9. After sequential staining of all markers, spectral DAPI (Akoya Biosciences, Marlborough, MA, USA) is then applied as the nuclear counterstain. Lastly, slides were mounted with ProLong Diamond Anti-fade Mountant (Molecular Probes, Life Technologies, USA) and cured in the dark at room temperature for 24 hours.

Images were acquired at 200X magnification using PhenoImager HT (Akoya Biosciences, Marlborough, MA, USA). Image quality checks were performed by checking staining positivity against known external control tissues positive for each marker and ROI were selected by pathologist (JY) using inForm v3.0 (Akoya Biosciences, Marlborough, MA, USA). HALO v3.6 (Indica Labs, Albuquerque, NM, USA) software was then used to perform cell segmentation. Thereafter, positivity (or negativity) of the marker was reduced to a binary classification by a threshold value determined. Finally, cell phenotyping was performed in HALO wherein combinations of markers were used to determine cell identity.

Cell–Cell Interaction Analysis in mIHC Data

The spatial relationship between various CD68⁺ macrophage phenotypes and tumor cells was quantified within individual ROIs using the average_percentage_of_cells_within_radius() function from the SPIAT package (v1.0.4). This function calculates the average percentage of CD68⁺ cells located within a 20 µm radius of tumor cells. Tumor cells were inferred from baseline samples in Panel 2 by excluding immune cell markers; specifically, cells negative for CD45, CD8, and CD68 were classified as tumor cells. In contrast, tumor cell identification was not feasible in Panel 1 due to the absence of the pan-leukocyte marker CD45, which is necessary for confidently excluding immune cell populations.

To assess the association between the spatial relationship of macrophage phenotypes and tumor cells with treatment response groups (PR vs. PD, PR vs. SD, and SD vs. PD), linear mixed-effects models (LMMs) were fitted using the lmer() function from the lme4 package (v1.1-35.1). P-values were computed using the lmerTest package (v3.1-3). To account for inter-sample variability due to multiple ROIs originating from the same tissue specimen, Sample ID was included as a random effect in all models.

Pathologist-Guided Tissue Category Annotation and Mapping to Visium Spots

Tissue categories were annotated on high-resolution H&E images by a pathologist (JV) using QuPath (v0.3.2). Annotated categories included stromal fibrosis, tumor epithelium, necrosis, non-neoplastic tissue, and several uncertain classifications due to suboptimal image quality: uncertain (favor neoplastic), uncertain (favor non-neoplastic), and uncertain. A custom Groovy script was used to export tissue category masks in PNG format from QuPath. A Python script was then employed to map Visium spot coordinates (as defined in tissue_positions.csv) to these tissue masks, which were scaled to match the H&E image resolution used in 10x Loupe Browser and spaceranger pipelines. For tissue category–specific analysis, Visium spots initially assigned as “uncertain” but expressing the hepatocellular carcinoma (HCC) marker GPC3 were reclassified as tumor epithelium for downstream analysis.

Pre-processing, Quality Control, and Clustering of Spatial Transcriptomics Data

For Visium, spatial transcriptomics data generated using the Visium platform were pre-processed with Space Ranger (10x Genomics), which aligned sequencing reads to the human reference genome refdata-gex-GRCh38-2020-A, assigned reads to spatial barcodes, and mapped gene expression to tissue spots based on H&E image alignment and fiducial frame detection, as visualized using Loupe Browser (10x Genomics). Following pre-processing, gene expression matrices from individual tissue samples were imported into Seurat (v4.4.0) for downstream quality control, analysis, and visualization. Quality control involved filtering out spots with fewer than 20 unique molecular identifiers (UMIs). Data normalization was performed using the SCTransform method, which accounts for technical noise and stabilizes variance across features. For each tissue sample, the top 3,000 most variable genes identified by SCTransform were used for downstream analyses, except in cases where raw counts were explicitly required.

For GeoMx DSP, spatial transcriptomics data generated using the NanoString GeoMx Digital Spatial Profiler (DSP) platform were processed using the GeoMx NGS Pipeline, GeoMxTools (v3.2.0), and standR[11] (v1.3.9). Sequencing reads in FASTQ format were converted into Digital Count Conversion (DCC) files using the geomxngs_fastq_to_dcc function. These DCC files were then aligned and quantified based on barcoded probe information and mapped to predefined gene panels (h.all.v2023.2.Hs.symbols.gmt) corresponding to each region of interest (ROI) or areas of illumination (AOIs a.k.a segments), using the geomxngs_dcc_to_count_matrix function. This process resulted in expression count matrices for a total of 260 ROIs/AOIs.

Quality control filtering was performed using GeoMxTools, with the default segment/AOI and probe QC parameters (removal of 47 segments) along with further filtering based on the detection rate of probe (%segments with detection) and segment (probe detection rate in each segment) at threshold 0.05 (or 5%) at LOQ=2 (removal of 12 segments). The count data from the filtered segments were normalized using the Trimmed Mean of M-values (TMM) method implemented in standR via the geomxNorm function. To account for technical variation, particularly slide-specific effects, the voom transformation followed by lmFit from the limma package was applied.

Tumor Region–Enriched Transcriptional Pathways

As an orthogonal approach to hdWGCNA, SPATA2 (v3.1.2)[12] was employed to identify gene programs enriched in tumor regions, using all other tissue regions within each Visium sample as the reference background. Samples lacking a visible tumor region, based on pathologist annotations, were excluded from the analysis. For each Visium sample, the spatial assay was first normalized and scaled using Seurat’s NormalizeData and ScaleData functions. Subsequently, a spataObject was initialized using the sample-specific count matrix, spatial coordinates, image scale factors, and with the spatial method set to "VisiumSmall". The normalized and scaled expression matrix was then added to the spataObject using the addProcessedMatrix function. To enable tumor region–specific differential expression analysis, tissue category annotations (i.e., “Tumor” or “other”) for each spatial coordinate were incorporated into the metadata of the spataObject. Differential expression analysis was then performed using the runDEA function with the method set to "wilcox". Tumor-enriched genes were extracted using the getDeaGenes function, specifying the target region as "Tumor" within the "Tissue" annotation column, using the input parameters across_subset = "Tumor" and across = "Tissue".

Hierarchical Gene Ontology Enrichment Analysis

To characterize the biological processes associated with each SM (generated by hdWGCNA; Fig. S7A-D)—represented by module eigengenes in Visium data—or with AOI-specific DE genes in DSP data, hierarchical Gene Ontology (GO) enrichment analysis was performed using topGO (v2.50.0) with human gene annotation and gene symbols (Fig. S7E-F). Fisher’s exact test was conducted via the resultFisher function to identify the top five GO terms significantly enriched for each module. GO terms were manually curated from the GO hierarchy: when multiple child terms contributed to a broader parent term, the higher-level parent term was recorded—unless the parent term was a root or immediate-child level category (e.g., “cellular process” or “metabolic process”), in which case the more specific child term was retained.

Word Cloud Visualization

To summarize gene programs associated with each SM—represented by module eigengenes in Visium data—or with AOI-specific DE genes in DSP data, word cloud visualizations were generated using the wordcloud (v2.6) and tm (v0.7-16) packages in R. GO term descriptions were first converted into a text corpus using VCorpus from the tm package. Text preprocessing was performed using tm_map, including: conversion to lowercase, removal of punctuation and numbers, elimination of common English stopwords (e.g., “the”, “and”), and stripping of extra whitespace. The cleaned corpus was then converted into a term-document matrix using TermDocumentMatrix, where rows represent terms (words) and columns represent tissue-specific SMs. Each matrix entry reflects the frequency of a given word in a particular SM. Word clouds were generated using the wordcloud package, with the following parameters: words were scaled by frequency, only terms with a frequency ≥ 1 were included, and a maximum of 100 words was displayed per cloud. Words were color-coded as either blue (low frequency) or red (high frequency).

Single Cell-type Differential Expression Analysis in GeoMx DSP

For GeoMx DSP, differential expression (DE) analysis was performed using edgeR on TMM-normalized counts, separately for CD8⁺ and CD68⁺ AOIs. The data were first converted into a DGEList object using SE2DGEList function, incorporating normalization factors computed via the TMM method. To model gene expression differences associated with treatment response, a linear model was constructed to assess pairwise comparisons between response groups (PR vs PD, PR vs SD, and SD vs PD), while controlling for slide-to-slide variation as a batch effect. The analysis followed the standard limma-voom pipeline with precision weights estimated using voom, and model fitting performed with lmFit.

Computational Identification of Spatial Niches Using Unsupervised Algorithms

Three deep learning models—stDCL (v1.0.1), Proust (latest version as of August 2025), and GraphST (v1.1.1)—were applied to identify spatial niches using Visium spatial transcriptomics data. Proust utilizes a graph convolutional autoencoder that integrates gene expression data with co-registered H&E images [13]. stDCL similarly employs a graph convolutional autoencoder, but operates on gene expression data alone [14]. In contrast, GraphST uses a non-graph-based autoencoder architecture, also relying solely on gene expression input [15]. For this purpose, Visium gene expression data—comprising spot coordinates and gene counts per spot—were first formatted into an AnnData [16][17] object to serve as model input. Default parameters were used for all models, except that the neighborhood radius in both Proust and GraphST was set to 5, reflecting the modest tissue area. In addition, the numbers of principal components for gene expression and image features in Proust were set to 50 and 10, respectively. To identify spatial niches, latent features generated by the three models—representing low-dimensional embeddings that capture spatial and transcriptional structure—were subjected to unsupervised clustering using Mclust (v6.1.1). Based on the five anticipated tissue categories (non-neoplastic, tumor epithelium, stroma, necrosis, and uncertain/others), the number of clusters was set to five for all three models during Mclust (v6.1.1) clustering, with default settings applied [18].

Bulk RNA-sequencing–derived HCC Biomarker Analysis for Distinguishing PD and SD

To evaluate the utility of previously reported HCC biomarkers [2]—originally shown to distinguish responders from non-responders—in discriminating more granular treatment response groups, particularly PD from SD within the non-responder population, we performed two complementary analyses. First, bulk RNA-sequencing–derived data were re-stratified into (PD, SD, and PR groups. Differences in pathway enrichment scores across response groups were assessed using the Kruskal–Wallis test, while associations with categorical variables—including chromosome 16 deletion status, immune subtype, and NCORI mutation status—were evaluated using Fisher’s exact test.

Second, HCC biomarkers previously identified from bulk RNA-sequencing analyses—including pathway- and gene-signature–based enrichment scores [2]—were re-computed using Visium spatial transcriptomics data and stratified into PD, SD, and PR groups. Enrichment scores were calculated using the fgsea function from the fgsea R package (v1.24.0). This included: (i) curated and Hallmark pathways from MSigDB (msigdbr v7.5.1; categories “C2” and “H”); (ii) the Kaya signature, an Asian-specific HCC gene signature; (iii) immune-related markers identified in an HCC immunotherapy cohort reported by Sangro et al. [19]; (iv) radiotherapy response–associated gene signatures (Rad) described by Dai et al.[20]; and (v) a nine-gene WNT/β-catenin pathway signature (WNT) reported by Grasso et al [21].

To assess the association between biomarker enrichment scores and treatment response (PR vs. PD, PR vs. SD, and SD vs. PD) at baseline and on-treatment time points, linear mixed-effects models (LMMs) were fitted using the lmer() function from the lme4 package (v1.1-35.1). P-values were obtained using the lmerTest package (v3.1-3). Sample ID was included as a random effect to account for within-response group variability. In addition, similar LMM analyses were performed to evaluate treatment-induced changes in biomarker scores (on-treatment vs. baseline) within each response group.

Cell Type Annotation for Spatial Transcriptomics Data

Cell type annotation based on gene marker enrichment was performed using MCPcounter (v1.2.0), an established transcriptome-based method for quantifying the relative abundance of immune and stromal cell populations in bulk and spatial transcriptomics data[22]. Specifically, SCTransformed expression matrices from Visium and TMM-normalized count data from DSP’s General ROIs were used as input. MCPcounter.estimate function computes enrichment scores for predefined cell types based on robust marker gene expression [22], enabling spatial inference of immune and stromal cell distributions across Visium spots and DSP ROIs.

In parallel, cell type annotation was performed via gene expression deconvolution using SPOTlight (v1.8.0), a method specifically designed for resolving multi-cell spot data in spatial transcriptomics platforms such as Visium [23]. This analysis leveraged a reference single-cell transcriptome to infer the immune and stromal cell type composition of each spatial location i.e. spot. To ensure relevance to our application, we trained a SPOTlight model using the publicly available single-cell Tumor Immune Cell Atlas as the reference dataset for cell type deconvolution. This atlas comprises approximately 500,000 cells from 217 patients across 13 cancer types[24] and is available as a Seurat object (TICAtlas.rds) from Zenodo (Record ID: 5205544).

The reference dataset was processed according to the recommended steps in the SPOTlight pipeline. First, the Seurat object was converted into a SingleCellExperiment (sce) object and log-normalized. Variance modeling was then performed using the modelGeneVar function, and the top 3,000 highly variable genes were selected using the getTopHVGs function. Cell type–specific marker genes were identified using the scoreMarkers function, based on the 30 level-2 cell types defined in the Tumor Immune Cell Atlas. Marker gene selection was guided by the area under the curve (AUC) scores returned by scoreMarkers, applying a threshold of AUC > 0.7 which was empirically determined. The resulting set of highly variable genes and cell type–specific marker genes, together with the query Visium gene expression data, were then provided as input to the SPOTlight() function. The output was a deconvolution matrix representing the estimated relative abundance of each cell type within each Visium spot.

Analysis of Spot-Wise Gene Expression in Visium

To quantify the spatial localization of immune cells and their potential interactions, the proportion of expressing spots (%spots) was calculated for selected single genes (CD8, CD68) and gene pairs (PDCD1–CD8, SPP1–CD68, and POSTN–CAF). For each tissue category, %spots were defined as the number of Visium spots with raw gene counts > 1 for the gene or gene pair (i.e. both target genes) of interest, divided by the total number of spots within that category.

Differences in gene expression abundance across cell types were evaluated using the non-parametric Kruskal–Wallis test.

Pre-processing and Resolution Optimization for Low-Dimensional Embedding–Based Gene Marker Identification

For Visium data, the 10,000 most variable genes across the 10 baseline tissue samples were identified by first merging the individual Seurat objects and applying log-normalization. SCTransform was then re-applied separately to each individual Seurat object, using the full set of available genes to generate normalized expression values. These SCTransform values were subsequently subset to retain only the globally defined top 10,000 variable genes. Among these, 1035 genes that were consistently detected across the Visium baseline samples were selected for downstream dimensionality reduction analysis. For DSP analysis, 23 ROIs from the baseline General ROI set were included. From the batch-corrected, TMM-normalized gene expression matrix, genes with a coefficient of variation (CV) greater than 25% were used for the dimensional reduction analysis.

To evaluate the appropriate resolution for identifying gene markers, unsupervised principal component analysis (PCA) was first performed on the pre-processed Visium and DSP datasets using the pca function from the mixOmics package (6.22.0). Although PCA of DSP data showed partial separation of patients by response group, PCA of Visium had limited clustering, indicating higher inter-sample heterogeneity and lower capacity to distinguish response groups based on spatial transcriptomic profiles alone (**Fig. S8A-B**).

Subsequently, supervised analysis was conducted using partial least squares discriminant analysis (PLS-DA). An initial PLS-DA model with 10 components was constructed for each dataset, and the optimal number of components was assessed using the perf function, which evaluates classification performance across components. Based on the resulting performance plots (classification accuracy vs. number of components), two components were selected for Visium, and three components for DSP, as they provided the best trade-off between accuracy and model simplicity. Contrasting to PCA, PLS of DSP data more effectively captured transcriptomic differences across response groups, suggesting its superiority in capturing subtle group-associated variations (**Fig. S8C-E**).

Signature Gene Identification and Importance Scoring

To identify gene markers associated with specific treatment response groups, a multi-step strategy was employed as follows. First, a heatmap of the 105 sPLS-DA–derived gene markers were generated and subjected to both gene-wise and sample-wise hierarchical clustering. Cluster assignments were defined using the cutree() function from the dendextend R package with *k = 4*, which achieved clear separation of the three known response groups. Genes that were uniquely enriched within each cluster corresponding to a response group were then identified. To quantify the discriminative power of each gene, an outcome-discrimination score was computed. This score was defined as the weighted sum of the gene’s absolute loadings from sPLS-DA, with weights corresponding to the proportion of response variance explained by each component.
