## Supplementary Figures for "Spatial Analysis Uncovers Immune Resistance Mechanisms in Non-Beneficial Hepatocellular Carcinoma Treated with Y90 Radioembolization-Nivolumab"

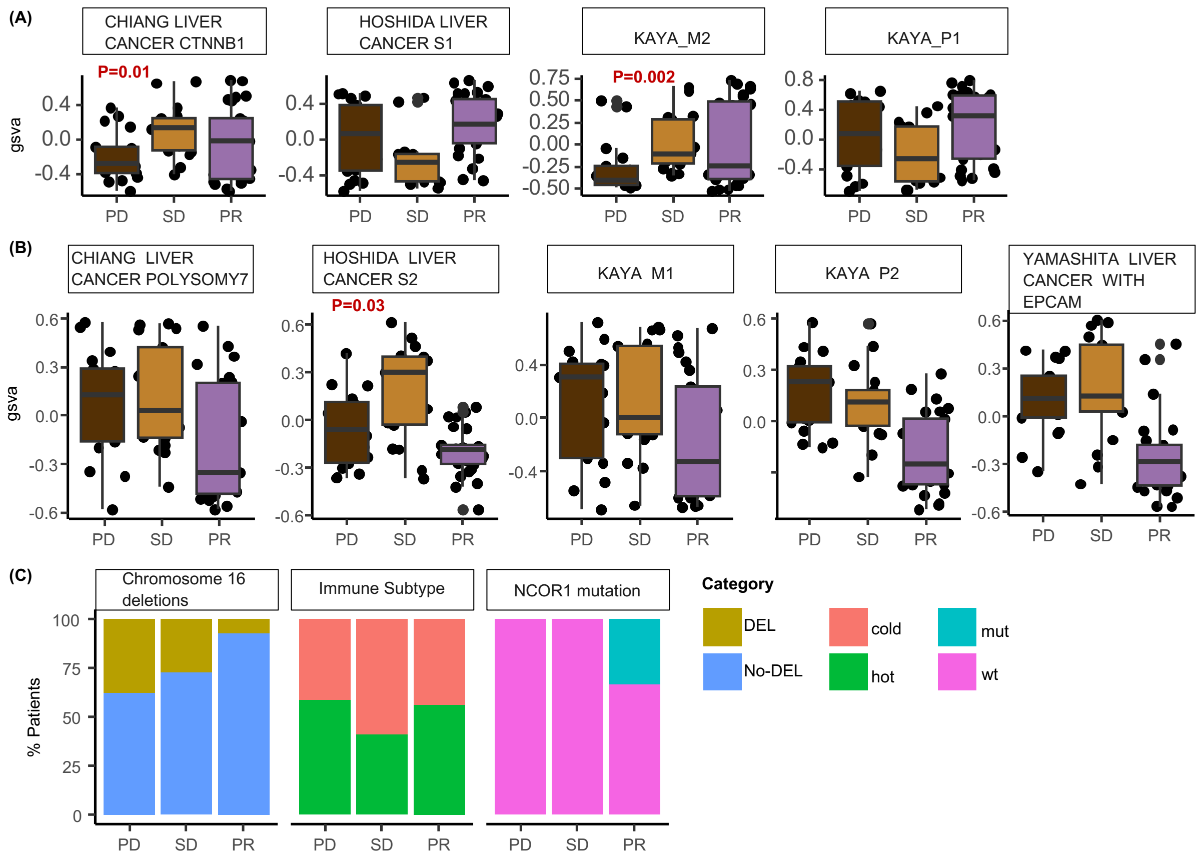


**Supplementary Figure 1. Baseline stratification by best clinical response (PD, SD, PR) to evaluate differential enrichment of HCC-related signature pathways and genomic/genetic biomarkers previously identified using bulk RNA-seq.** (A-B) GSVA-derived enrichment scores for pathways upregulated in (A) responders (i.e. PR) and (B) non-responders (including PD and SD), respectively.(C) Relative distribution of patients based on binary classification of genomic/genetic biomarkers, stratified by best response groups: PD, SD, and PR. Statistical significance was assessed using the Kruskal–Wallis test for (A–B) and Fisher's exact test for (C) to examine associations between PD and SD (within the responder group) and individual pathways or genomic/genetic biomarkers; P-values >0.05 were not shown.

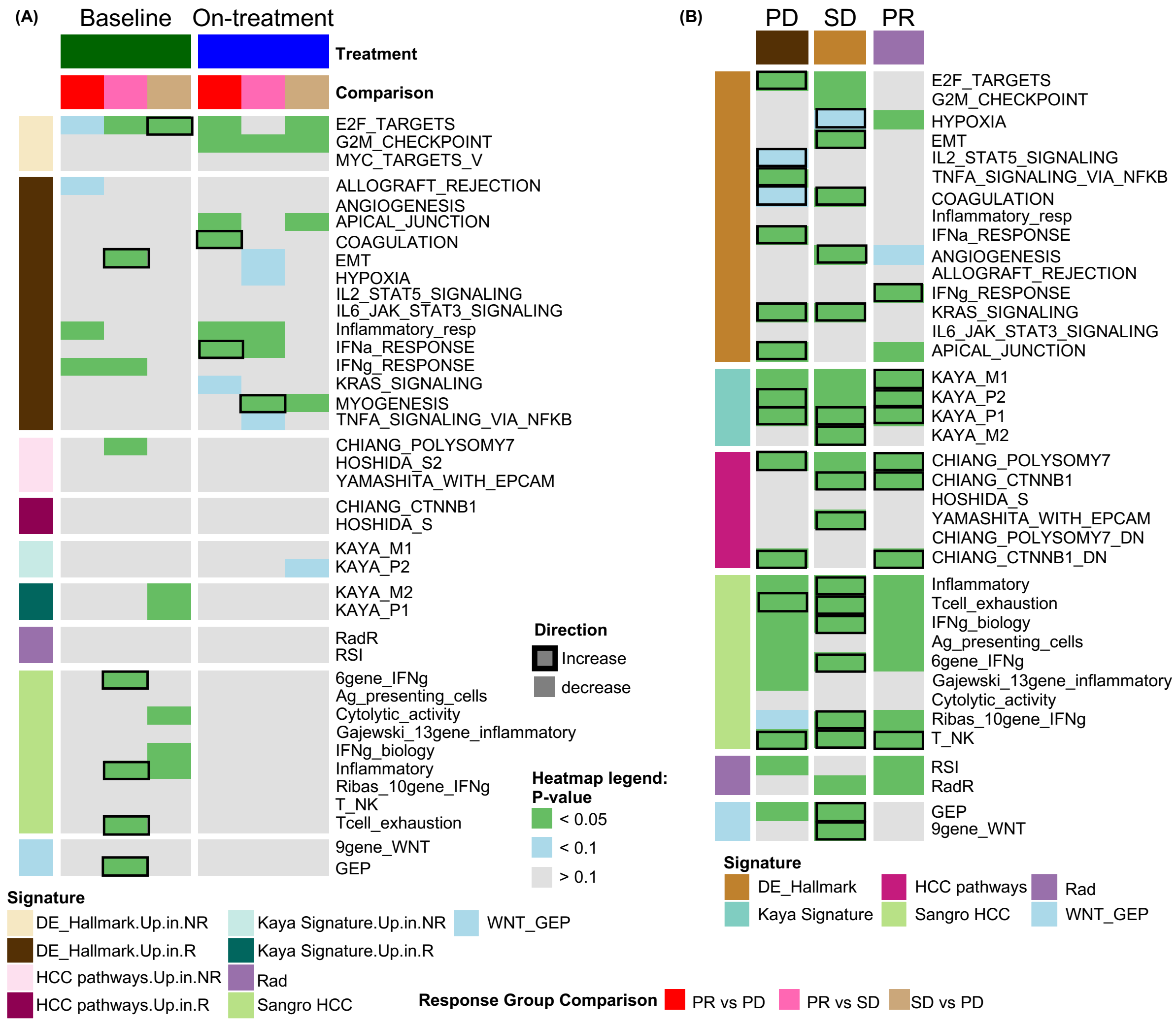


**Supplementary Figure 2. Spatial enrichment of HCC-related signature pathways, derived from literature or previously identified through bulk RNA-seq, analyzed using Visium spatial transcriptomics.** (A) Differences in pathway enrichment scores across treatment response groups (PD vs. SD, PR vs. SD, and PR vs. PD). (B) Differences in pathway enrichment scores between on-treatment and baseline samples. Statistical significance was assessed using a linear mixed-effects model adjusting for tissue effects. Pathway definitions: DE_Hallmark: Differentially expressed hallmark pathways identified from bulk RNA-seq (Kaya et al.); Kaya Signature: Asian-specific HCC gene signature from Kaya et al.; HCC Pathways: Markers reported in an HCC immunotherapy cohort (Sangro et al.); Rad:

Radiotherapy response-related gene signatures (Dai et al.); WNT: A nine-gene Wnt/β-catenin pathway signature (Grasso et al.).

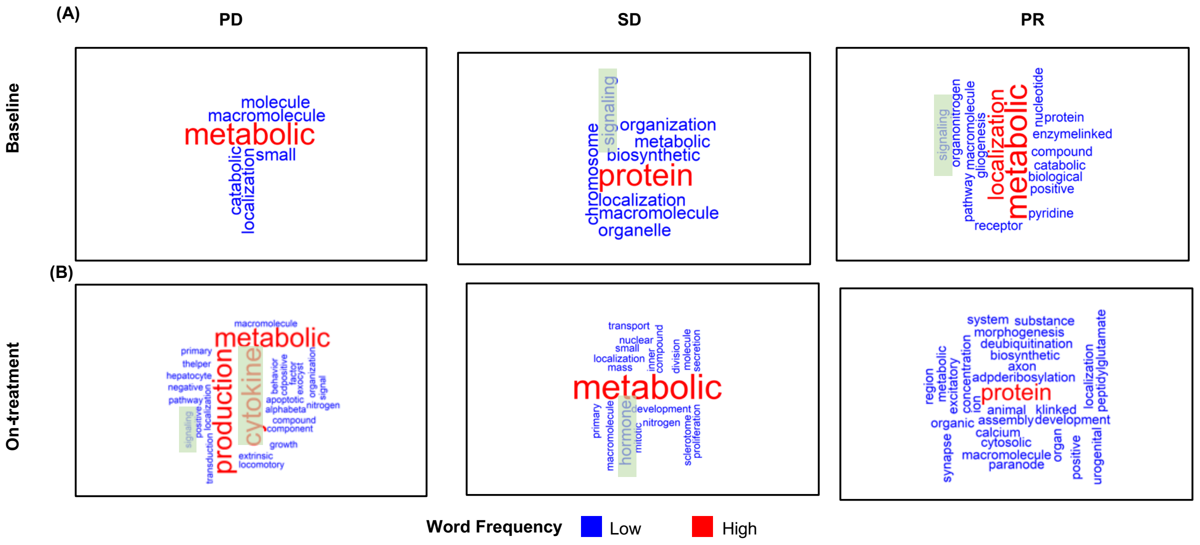


**Supplementary Figure 3. Tumor region–enriched gene programs identified using SPATA2 and hierarchical GO analysis in Visium data.** Differential expression analysis was performed using SPATA2, comparing gene expression between pathologist-annotated tumor regions and surrounding non-tumor regions within each Visium sample. Enriched gene programs within tumor regions were subsequently identified through hierarchical GO analysis. GO terms were consolidated by treatment response group: (A) baseline and (B) on-treatment. Results were visualized as word cloud plots. Immune-related GO terms are highlighted in green boxes. Note: Visium samples consisting solely of tumor regions were excluded from this analysis due to the absence of non-tumor reference regions—a current limitation of SPATA2.

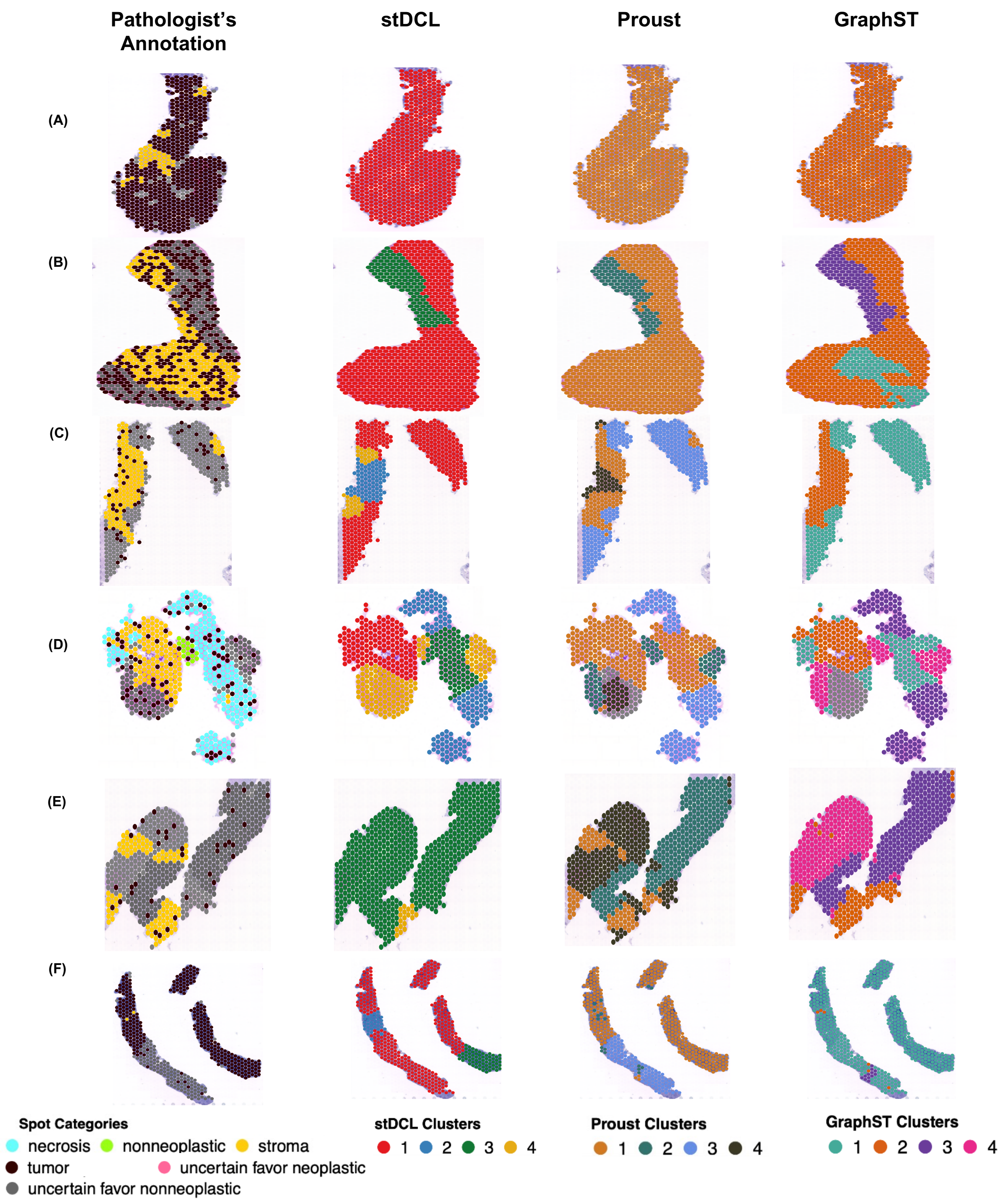


**Supplementary Figure 4. Comparison of pathologist-annotated tissue categories (leftmost column) with unsupervised clustering results from three multi-omics spatial clustering algorithms — stDCL, Proust, and GraphST (second to fourth columns) — across five representative Visium samples (A-F).**

No consistent alignment with the pathologist-defined ground truth is observed for any of the three unsupervised multimodal spatial transcriptomics clustering methods.

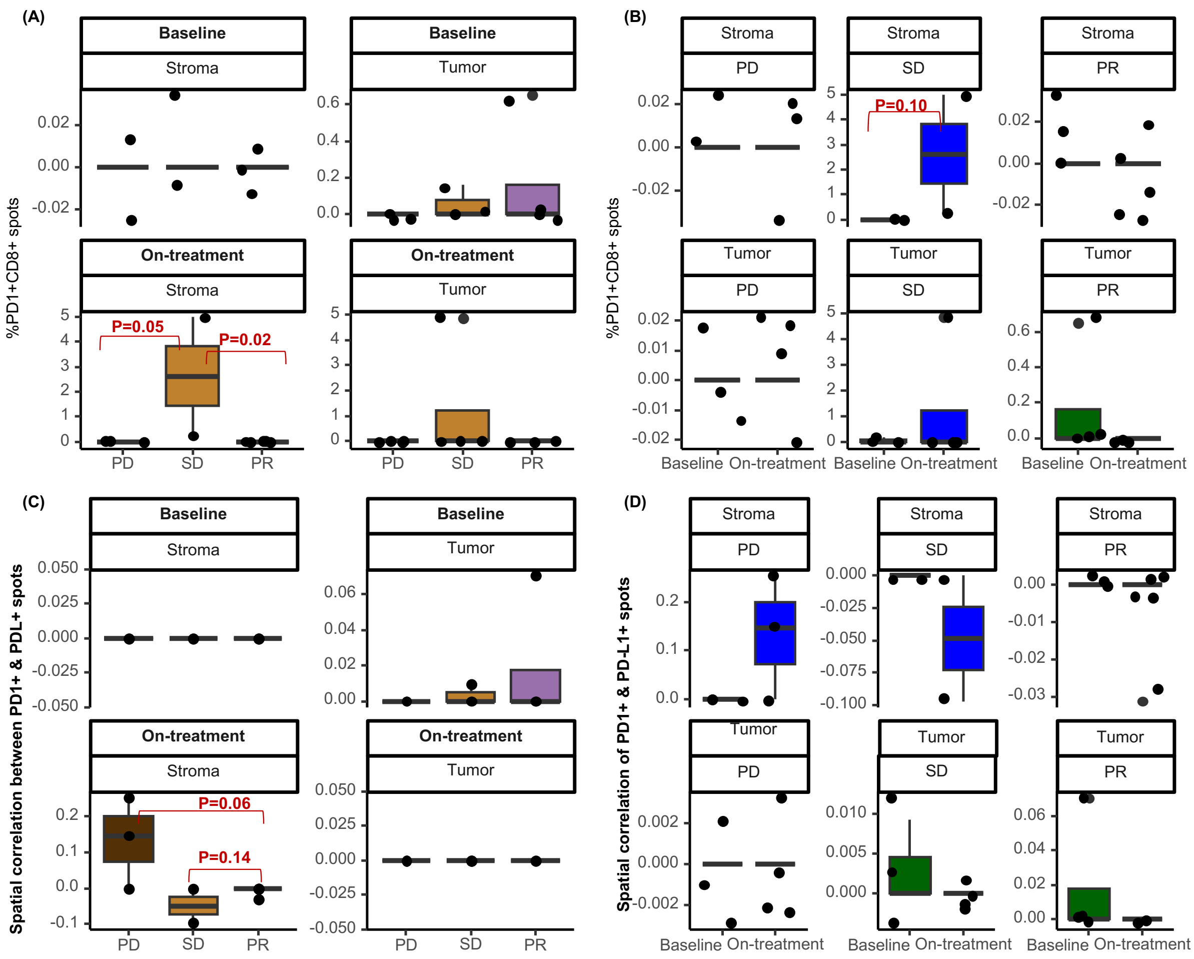


**Supplementary Figure 5. Spatial relationship and abundance of CD8, PD1, and PDL1 assessed using Visium.** (A–B) Comparison of CD8–PD1 colocalization (percentage of spots) between response groups: (A) PR vs SD, SD vs PD, and PR vs PD; (B) baseline vs on-treatment. (C–D) Comparison of spatial correlation between PD1 and PDL1 (Spearman correlation across neighboring spots): (C) across response groups—PR vs SD, SD vs PD, and PR vs PD; (D) baseline vs on-treatment. P-values > 0.1 are not shown.

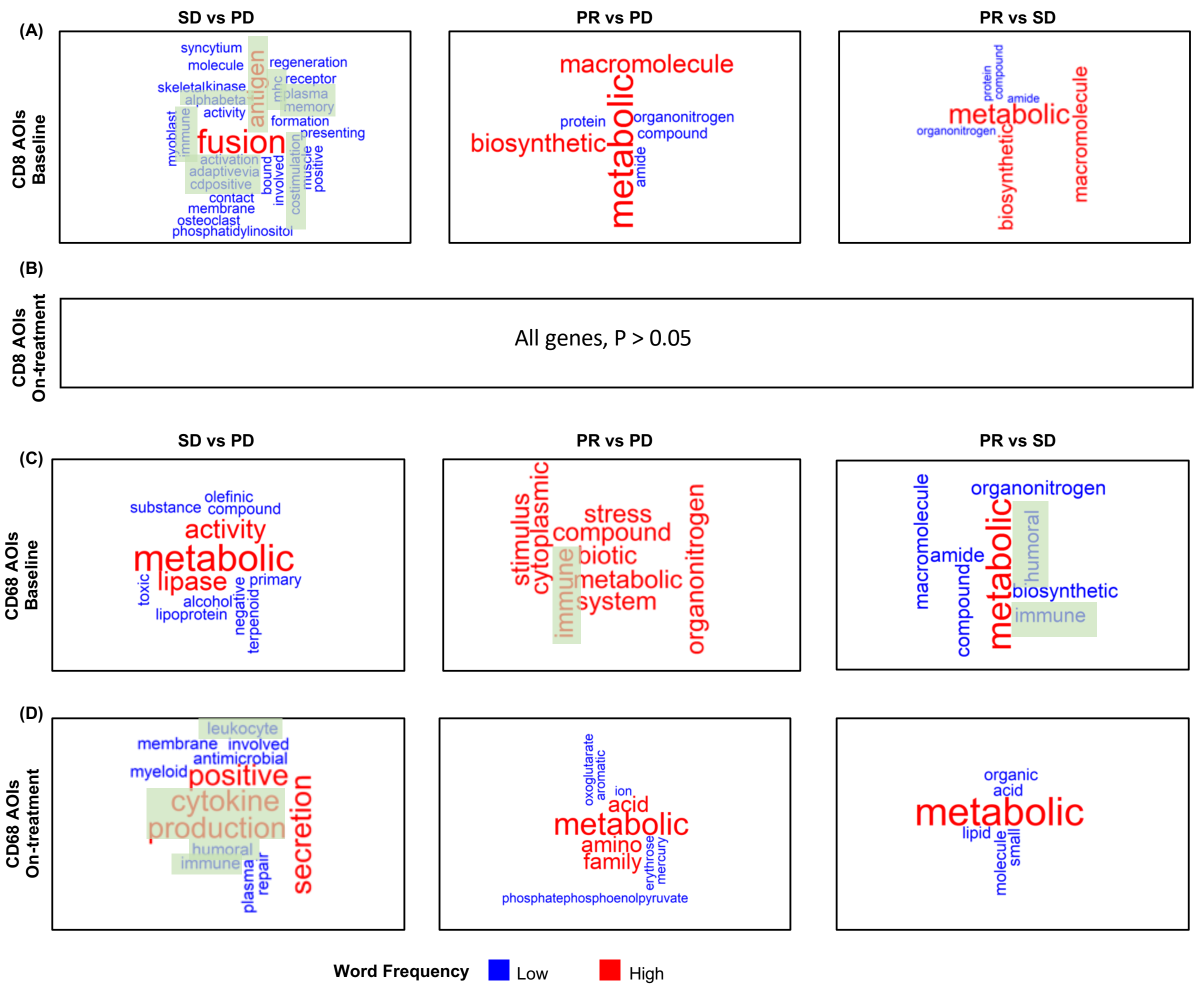


**Supplementary Figure 6. Single cell-type differential gene expression and GO analysis using CD8$^+$ and CD68$^+$ AOIs in DSP data at baseline and on-treatment.** Differentially upregulated genes were identified from: (A) CD8$^+$ AOIs at baseline: comparisons from left to right—SD vs PD, PR vs PD, and PR vs SD, (B) CD8$^+$ AOIs on-treatment: no significantly differentially expressed genes were detected; (C) CD68$^+$ AOIs at baseline: comparisons from left to right—SD vs PD, PR vs PD, and PR vs SD; (D) CD68$^+$ AOIs on-treatment: comparisons from left to right—SD vs PD, PR vs PD, and PR vs SD. Genes from each analysis were subjected to hierarchical GO analysis, and the resulting GO terms were visualized using word cloud plots. Words extracted from immune-related GO terms were highlighted in green boxes.

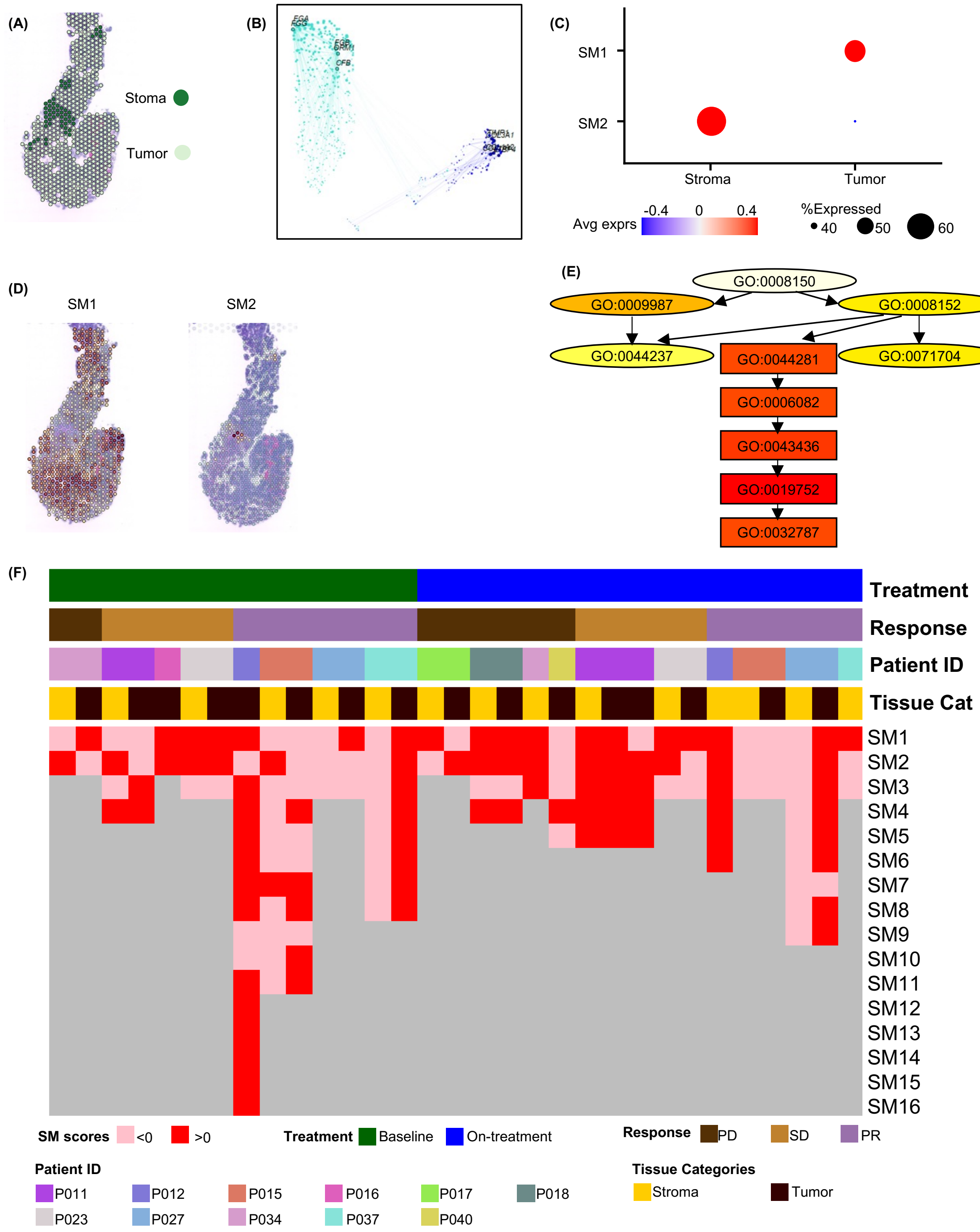


**Supplementary Figure 7. Illustrative example of hdWGCNA analysis and summary heatmap of spatial module eigengenes across all samples.** (A–E) Example using a representative tissue sample: (A) Pathologist-annotated tissue regions; (B) Gene co-expression network visualized in UMAP embedding; (C-D)

Spatial modules (SMs) and their corresponding module eigengene expression scores shown as (C) dot plots grouped by stromal fibrosis and tumor regions, and (D) spatial maps overlaid on tissue sections; (E) Hierarchical GO analysis using eigengenes of SM1, with the five most specific (deepest) GO terms used to generate word clouds (see Fig. 1C). (F) Heatmap displaying average SM eigengene expression (binarized as >0 or <0), with columns ordered by treatment timepoint, response group, sample ID, and tumor/stroma classification for all Visium samples.

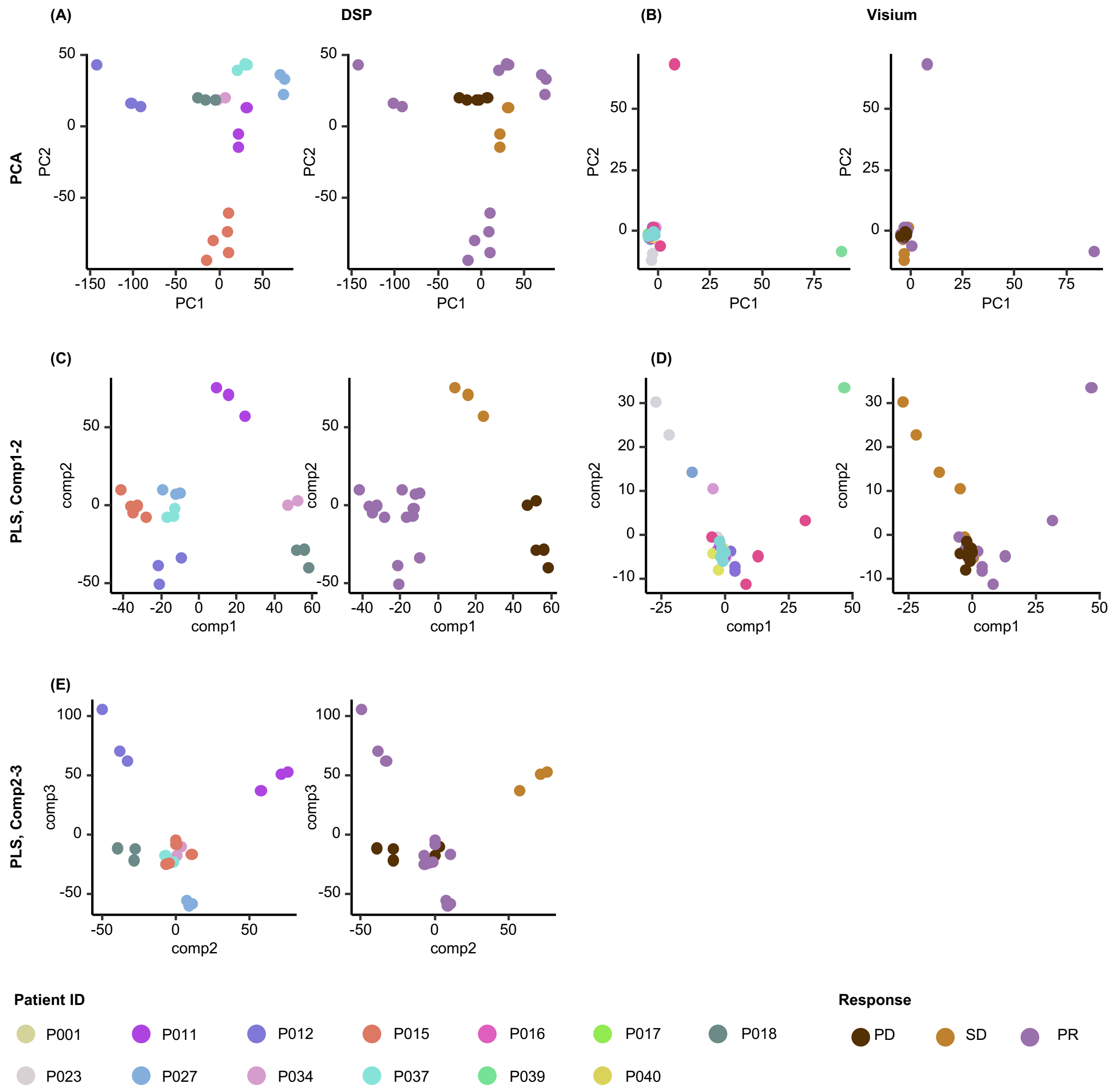


**Supplementary Figure 8. Dimensionality reduction analysis using PCA and PLS on DSP and Visium data.** (A, C, E) Sample distribution using DSP data: (A) PCA space; (C–E) PLS space—each panel shows samples annotated by patient ID (left) and response group (right). (B, D) Sample distribution using Visium data: (B) PCA space; (D–E) PLS space—each panel shows samples annotated by patient ID (left) and response group (right).
